# An Evaluation of the Relationship Between Public Communications on Food Recalls and Online Self-reporting of Foodborne Illness

**DOI:** 10.1101/2025.07.24.25332152

**Authors:** Yu Wang, Yu Cao, Hugh Rand, James Pettengill

## Abstract

The US Food and Drug Administration routinely announces product recalls to inform the public about mislabeled or potentially hazardous products. For example, food recalls may occur when a product is contaminated with a foodborne pathogen (e.g., *Listeria monocytogenes*) or contains an undeclared allergen (e.g., nuts). In addition to governmental systems that capture consumer or industry self-reported problems with food at the federal, state and county level, there are platforms such as https://iwaspoisoned.com (IWP) through which the public can self-report when they are sick, share their symptoms, and note what they think made them sick. Here we use the Granger Causality (GC) test to investigate two information streams – IWP and FDA recall announcements – to determine whether there is a statistically supported relationship between them and if so the extent of that relationship (i.e., to understand if self-reporting through IWP precedes recalls or vice versa). Among 48 foods selected due to their high number of occurrences in both IWP and FDA recalls, whether IWP was predictive of bacterial recalls had the highest number of positive GC tests with 15 foods (31%). There were 12 foods that had positive GC tests for both bacterial recalls being predictive of increased IWP reports and allergen recalls being predictive of increased IWP reports. The lack of a stronger relationship between the information streams does not detract from the importance and utility of self-reporting platforms and communications by public health agencies. Rather they illustrate that based on the method employed here that the influence on public behavior and the degree to which self-reporting can forecast future recalls is difficult to detect. However, with further developments in the modeling of the relationship, data capture approaches, and public communication strategies the relationship will likely be improved.

## Introduction

The United States Center for Disease Control and Prevention (CDC) estimates that each year in the United States there are 47.8 million episodes of illness, 127,839 hospitalizations, and 3,037 deaths due to consumption of contaminated foods [1]. As a result, numerous state, federal, industry, and other partners expend resources to minimize the risk. This includes the US Food and Drug Administration (FDA) coordinating with industry as well as with its federal, state, and local partners on food product recalls. Such recalls occur because of hazards that products present; for example, a recalled product could be contaminated with a foodborne microbial pathogen (e.g., *Escherichia coli* O157:H7; [2]) or contain an unnamed allergen (e.g., nuts; [3]) both of which can have dire health consequences.

In addition to state- and government-funded surveillance programs, in the age of social media and widespread internet access there are additional surveillance mechanisms that may be of use to public health practitioners. The utility of such online sources has greatly increased in the past few years due to developments in machine learning, text mining, and natural language processing (NLP). For example, supervised machine learning and text mining approaches have been applied to extract signals indicative of food poisoning or other adverse effects from online reviews on www.amazon.com and self-reports submitted to www.iwaspoisoned.com (IWP) [4]. Results were mixed where for top-ranking reviews the model was 77-90% accurate but sentiment was only 11-26% accurate; the authors also benchmarked their hazard-flagged products by consulting a panel of 21 food safety experts [4]. Another study [5] applied similar approaches to the large amount of data on the online social media platform Twitter (now known as X), but also used NLP and specifically the large language model BERTweet [6] to help extract signals related to food safety. In that study, they used the CDC’s NORS (National Outbreak Reporting System, https://www.cdc.gov/nors/index.html) data to benchmark model performance and found that in general the most implicated food categories and their distributions were similar to those provided by NORS [5].

Here we investigate the extent to which the signals in a self-reporting platform for food safety may be influenced by public communications on food recalls and whether signals provided by the self-reporting platform precede a food safety risk event that results in a recall. We focus on IWP, which is a consumer platform to improve food safety by sharing consumer experiences, and on FDA recalls. On the IWP platform consumers can report their suspected foodborne illness experiences which are shared with the public, health authorities, and food businesses.

Determining the correlation and direction of the relationship between IWP and public announcements on food recalls may help with understanding how to interpret signals from self-reporting platforms and how such platforms can be used in support of protecting the public health, and improving public communications about food safety.

## Materials and Methods

### Raw Data

Reports of potential food poisoning between Jan. 1, 2016 and Mar. 31, 2023 were downloaded from IWP (https://iwaspoisoned.com/); this provided 357,971 individual reports. Each report usually, but not always, corresponds to a single individual reporting one incident of gastrointestinal distress. Only reports with type “poison” were used in order to focus on food poisoning, which resulted in 329,709 reports. Data on FDA food recalls were downloaded from the FDA Enforcement Report website (https://www.accessdata.fda.gov/scripts/ires/) by selecting “Food” as the “Product Type”; this provided 3,682 individual food-related recall entries. Note that any situation requiring the issuing of a recall may have multiple entries corresponding to different lot codes or production dates. It is also important to note that we do not include recall announcements and data from other US public health agencies such as USDA FSIS (Food Safety Inspection Service).

### Food name extraction

To match food items between IWP and FDA recall data we used the FoodOn ontology (https://github.com/FoodOntology/foodon/blob/master/foodon-synonyms.tsv) to classify like products based on the complete food names. For each FDA recall entry, we searched words in the “Product Description” field for food names. These identified food names in FDA recall data were used to identify food names in IWP reports. The free-text “report title” and “body parts” fields in each IWP report were examined to find any words that also occurred in the food names found in FDA recall data. Because the fields contain free-text entries it is challenging to unambiguously determine the attributed food so we searched by food names identified in FDA recall reports to help mitigate this problem. To also account for the noisiness in these free-text fields, we applied a fuzzy matching method (package used: FuzzyWuzzy, https://pypi.org/project/fuzzywuzzy) that finds words with a similarity score of at least 85 out of 100 to a certain food followed by manual review. Checking of a subset of the categorized data provided an estimated error rate of <0.1%.

### FDA recall report categorization and food selection

Recall entries were split into categories based on contamination type. For each entry, if the text in column “Reason for Recall” contained “cronobacter”, “campylobacter”, “listeria”, “l. monocytogenes”, “salmonella”, “e. coli”, “e.coli”, or “Escherichia”, then the entry was classified as bacterial; otherwise, if the text contained any of the eight major foodborne allergens (https://www.fda.gov/food/buy-store-serve-safe-food/food-allergies-what-you-need-know), then the entry was classified as allergen. When neither of these categories were identified, the entry was discarded. The initial 3,682 entries provided 946 bacterial and 1,542 allergen recall entries, respectively. The total numbers of times that each food was mentioned in IWP reports and FDA bacterial and allergen recall reports were counted. Only food names mentioned more than 1,000 times in IWP reports and at least 10 times in either FDA bacterial or allergen recalls in this 7-year period were selected.

This resulted in 44 foods (Table 1).

**Table 1.** The Granger causalities summary between IWP reports and FDA recalls across 44 foods (see Figure 2 for more information)

|  | No | Yes |
| --- | --- | --- |
| IWP Granger cause allergen recalls | 33 | 11 |
| Allergen recalls Granger cause IWP | 34 | 10 |
| IWP Granger cause bacterial recalls | 31 | 13 |
| Bacterial recalls Granger cause IWP | 35 | 9 |

### Time series analysis

For each food, we generated three time series with a length of 2647 days (January 1, 2016 to March 31, 2023): one for IWP reports, and one each for FDA bacterial and allergen recall data. Each data point represented the number of records for the food mentioned for that day. The augmented Dickey-Fuller (ADF) test was performed to assess the stationarity of each time series (i.e., the statistical properties of a time series do not change over time); non-stationary time series were discarded. Considering consumption of a contaminated food batch occurs over a period of time, a moving-average (MA) was applied to IWP time series. Since we were not sure about the length of that period for different foods, we ADF-tested time series with MA window sizes from 1 day to 30 days. For each food with stationary time series, Granger Causality (GC) tests were performed to evaluate the predictive power between IWP reports and FDA recall events. Furthermore, when performing GC test, lags ranging from 1 day to 60 days were tested individually to find the possible predictive power. The ADF test and GC test were done with Python package “statsmodels” (https://www.statsmodels.org/stable/index.html). Figure 1 provides a diagram of the time series analysis steps.

**Figure 1.**
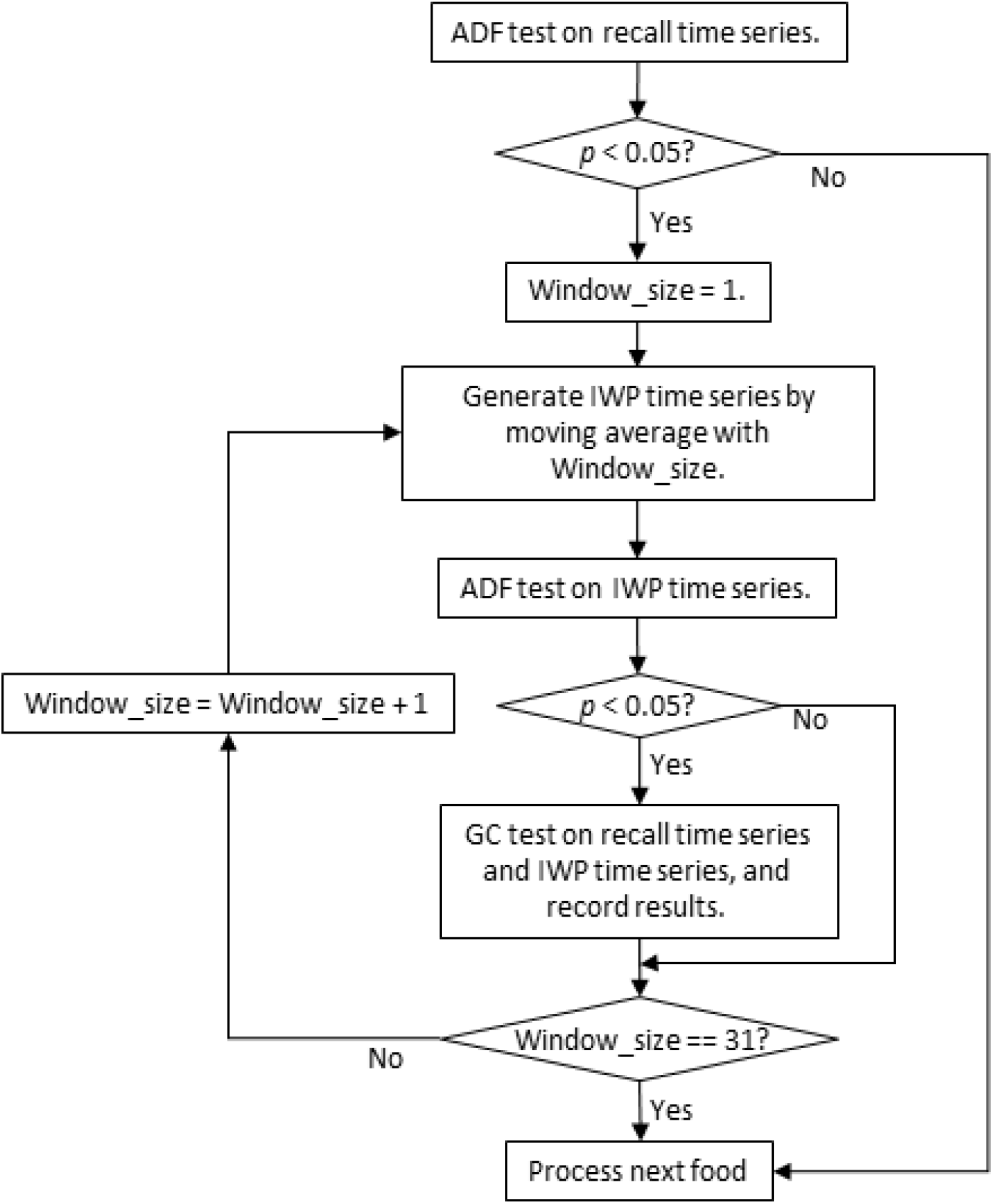
Steps of statistical analysis on time series of IWP reports and FDA recalls.

For each GC test the package returns four results: two based on the F distribution, and two on the chi-square distribution. The null hypothesis for a GC test (i.e., one time series does NOT predict the other time series) was rejected only when all four ***p***-value were less than 0.05. We discuss the use and interpretation of the results in the discussion section.

## Results and Discussion

The GC test that we applied in this study assesses whether one time series (the assumed “cause”) contains unique information that can be used to predict the future values of the other time series (the assumed “effect”) (8). Regardless of its name, it is not a test of a causal relationship. The standard test relies on the time series being stationary (not fluctuating); hence, we tested both the IWP time series (i.e., the ones with different window sizes for moving average) and the FDA recall time series with the ADF test. The test results showed that all FDA recall time series were stationary, while 232 out of 1,320 IWP time series were not and therefore removed from the GC tests.

The results of the GC tests for the 44 foods and their associated recall and IWP data identified via our prioritization could fall into 1 of 4 groups: IWP is predictive of allergen recalls; allergen recalls are predictive of IWP; IWP is predictive of bacterial recalls; and bacterial recalls are predictive of IWP. To minimize spurious significant results, for each group in each food, the result was “Yes” only when the GC test results of at least 10 different moving average window sizes in at least two lags were significant. Among the 4 groups, the highest number of positive GC tests was 13 out of 44 foods (30%) in the group where IWP is predictive of bacterial recalls (Table 1). However, the other GC tests for the directionality of a cause were somewhat similar and as a result there was no statistically meaningful difference in positive results among the possible relationships between IWP and allergen or bacterial food recalls (e.g., it is not more likely to see IWP self-reported data predict a recall than it is for a recall to elicit an increased IWP reporting rate). There were 19 foods with no predictive power between IWP reports and the two types of FDA recalls (Supplemental Table S1). For three fruits (apple, banana, and strawberry), no GC exists between IWP reports and FDA allergen recalls, while GC did exist between IWP reports being predictive of bacterial recalls for both apple and banana. For multi-ingredient food in this table (cake, cookie, pie, pizza, roll, salad, and sandwich), no GC exists except one for sandwich.

Though the GC test is misnamed and does not test the existence of real causality, it does provide information in the form of temporal correlation. For example, for “chips”, allergen recalls appear to elicit an uptick in IWP reporting with about a 10-day or more lag (Supplemental Figure S2). During the whole period of 2647 days, there are 78 allergen recalls for chips, with a peak on July 19—20, 2018 (Recall event IDs: 80574, 80576, 80587, and 80603). In IWP reports, the 7-day average count for chips is 1.05±0.52 (mean±std, max=2.28) within 60 days before July 19, 2018, and jumped to 2.85±2.47 (mean±std, max=7.43) in the following 4 weeks after July 20, 2018 (Figure 2). On the other hand, for foods with no positive GC test results between IWP reports and FDA recalls like chicken and cookie, we do not see such temporal correlation.

**Figure 2.**
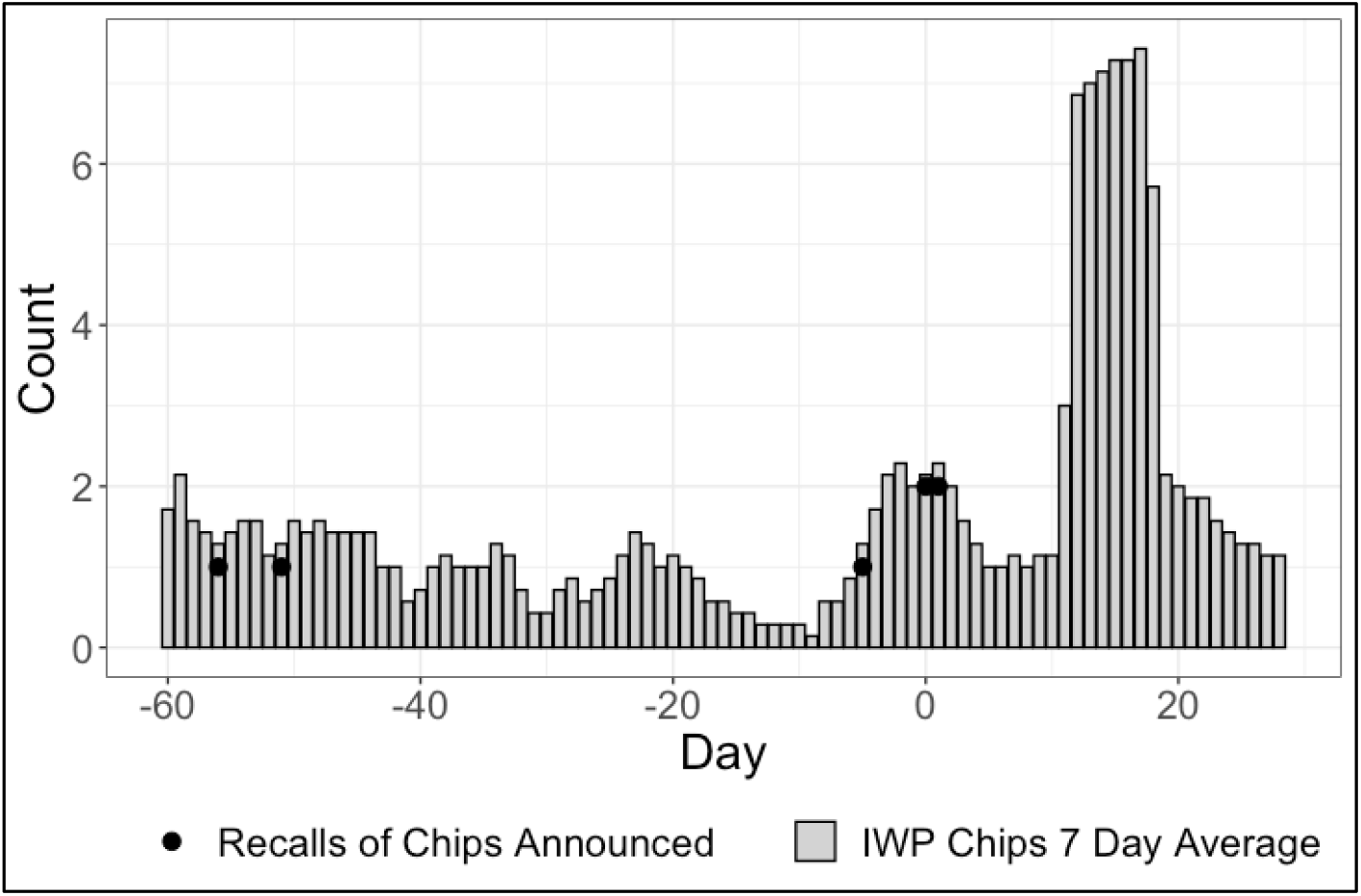
IWP 7-day average for chips before and after 4 FDA allergen recalls for chips. Day 0 is July 19, 2018.

Different factors may explain the results we observed, where only about a third of food types had some statistically supported relationship between recalls and IWP reporting. One potential explanation is that the public is not that influenced by recall data and, therefore, we would not expect to see an uptick in IWP reports following a recall announcement. There is some evidence to suggest this is the case given that public communications of foodborne illness in the form of outbreak announcements get the attention of the public more so than recalls, where the latter “may not generate social media posts” [7]. We have only focused on FDA recall data and not incorporated additional recall announcements from, for example, FSIS. As a result, there may be more of a relationship between the public communications from health agencies and self-reporting mechanisms like IWP than reported here but additional research is needed to confirm. There are also likely methodological difficulties that may inhibit detection or recognition of a signal. For example, IWP data is free-form unstructured text, and identification of a specific food type can be challenging because of the ambiguity of terms used, that multiple food types are mentioned, lack of standardization, and other reasons. The difficulties of working with unstructured free-text data within public health research are well-documented [8], but the emergence of foundational LLMs (Large Language Models) may help ameliorate some of those difficulties [9]. The FDA recall data is also not without difficulties, where some recalls of multi-ingredient foods did not provide a full list of ingredients. Another factor to consider with foodborne disease reporting is the time lag between exposure and disease onset, ranging from several days to weeks. Norovirus is an exception to this, but norovirus tends to be a local problem and very unlikely be present in FDA recall reports. Supporting this is the fact that only 9 of 3,682 recall events were due to norovirus contamination, despite the ranking of norovirus as the leading cause of foodborne illness in the United States. This makes consumer attribution of a specific food to a given case of gastrointestinal distress likely to be of low accuracy. Allergic reactions are generally much more immediate, and people with allergies generally know how they tend to react. This makes allergen problems readily apparent in the IWP data.

## Conclusions

Effectively addressing food safety requires integration of many different types of information from different sources. Our analysis indicates that for some food, IWP reports can be used to predict future FDA food recalls, though a predictive model with high accuracy may need to include other factors such as season, location, and food type in addition to more advanced unstructured text processing methods. The recalls announced by the FDA were also at times concordant with and preceded an increase of reports within that platform. Findings from our work in assessing the connection between known issues (i.e., recalls) and reported issues (i.e., IWP) indicate that while integrating information streams poses challenges, public platforms such as IWP may play an important role in evaluating the efficacy of public communications. Our work showing instances where IWP reporting may precede food recall events also provides additional support for the utility of such platforms for surveillance and their ability to help detect problems.

## Data Availability

All data produced are available online at http://www.iwaspoisoned.com and
https://www.accessdata.fda.gov/scripts/ires/

https://www.accessdata.fda.gov/scripts/ires/

## Acknowledgements

We thank P. Quaid of IWP for insightful discussions on the use and interpretation of IWP data. We also thank B. Wolpert for providing feedback on drafts of the manuscript.

