## Supplemental Table S1 for "An Evaluation of the Relationship Between Public Communications on Food Recalls and Online Self-reporting of Foodborne Illness"

Table S1. The Granger causalities (i.e., predictive power) between IWP reports and FDA recalls of 44 foods (see Figure S2 for more information)

| Food | IWP predictive of allergen recalls | Allergen recalls predictive of IWP | IWP predictive of bacterial recalls | Bacterial recalls predictive of IWP |
|---|---|---|---|---|
| apple | No | No | Yes | Yes |
| bacon | Yes | No | No | No |
| banana | No | No | Yes | No |
| bean | Yes | No | No | No |
| beef | No | No | No | No |
| bread | Yes | No | No | Yes |
| butter | No | Yes | Yes | Yes |
| cake | No | No | No | No |
| cheese | No | Yes | Yes | No |
| chicken | No | No | No | No |
| chili | No | No | No | No |
| chips | No | Yes | Yes | No |
| chocolate | No | No | Yes | No |
| coffee | No | No | No | No |
| cookie | No | No | No | No |
| corn | Yes | No | Yes | No |
| cream | Yes | Yes | Yes | No |
| dairy | No | No | No | No |
| egg | No | No | No | Yes |
| fish | Yes | No | No | No |
| garlic | No | No | Yes | No |
| honey | No | No | No | No |
| juice | No | No | No | No |
| lettuce | No | No | No | No |
| milk | Yes | No | Yes | Yes |
| mushroom | No | No | No | Yes |
| onion | No | Yes | Yes | Yes |
| pasta | No | No | No | No |
| pepper | No | No | No | No |
| pie | No | No | No | No |
| pizza | No | No | No | No |
| potato | No | Yes | No | No |
| rice | Yes | Yes | Yes | No |
| roll | No | No | No | No |
| salad | No | No | No | No |
| salmon | Yes | No | No | No |

| | | | | |
|---|---|---|---|---|
| sandwich | No | No | No | Yes |
| shrimp | No | Yes | No | Yes |
| spinach | No | No | No | No |
| strawberry | No | No | No | No |
| tea | Yes | No | No | No |
| tomato | Yes | Yes | Yes | No |
| tuna | No | Yes | No | No |
| turkey | No | No | No | No |
