## Supplemental Figure S2 for "An Evaluation of the Relationship Between Public Communications on Food Recalls and Online Self-reporting of Foodborne Illness"

Supplemental Figure S2. Results of Granger causality tests for 48 selected foods.

"allergen_ds_gc_recall": IWP predicts allergen recalls.

"allergen_recall_gc_ds": allergen recalls predicts IWP.

"bacterial_ds_gc_recall": IWP predicts bacterial recalls.

"bacterial_recall_gc_ds": bacterial recalls predicts IWP.

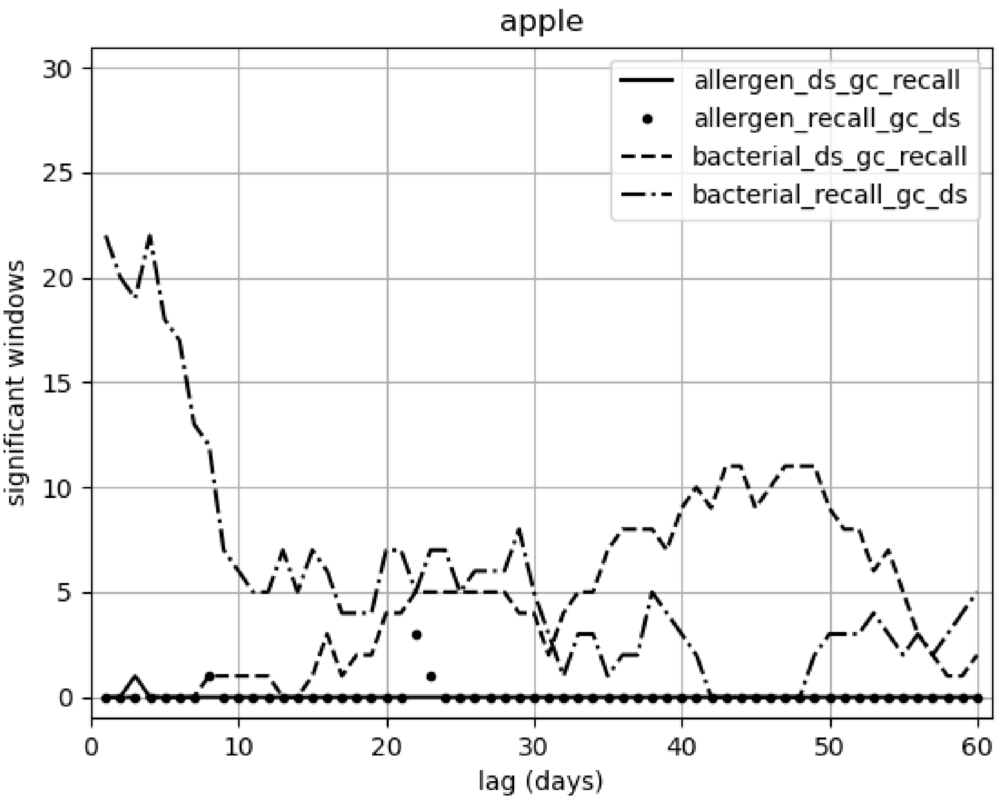

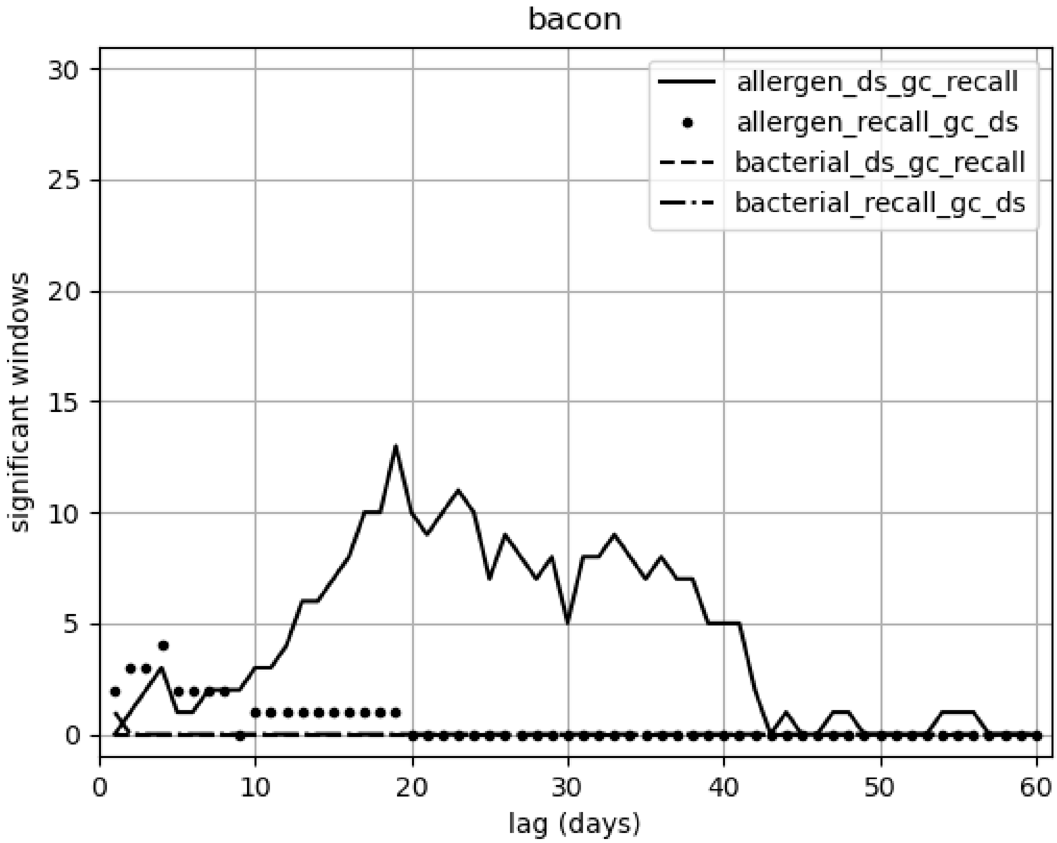

bacon
allergen_ds_gc_recall
allergen_recall_gc_ds
bacterial_ds_gc_recall
bacterial_recall_gc_ds
significant windows
lag (days)
0
5
10
15
20
25
30
0
10
20
30
40
50
60

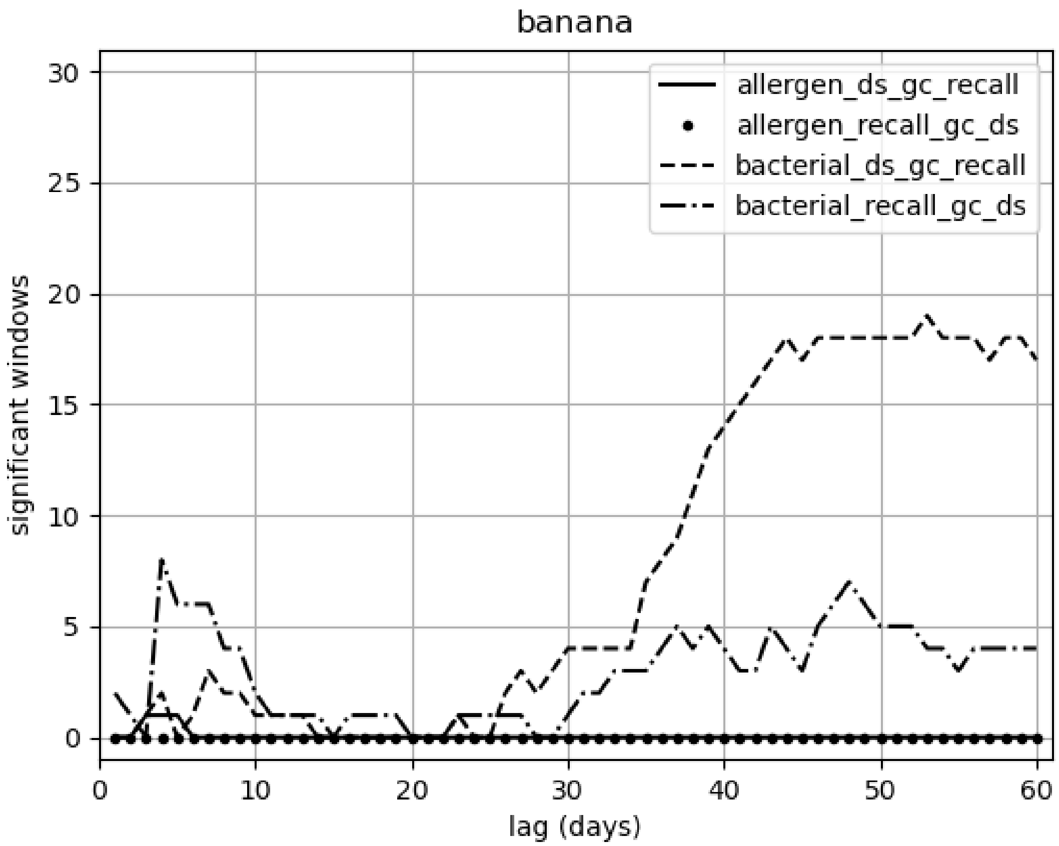

banana
allergen_ds_gc_recall
allergen_recall_gc_ds
bacterial_ds_gc_recall
bacterial_recall_gc_ds
significant windows
lag (days)
0
5
10
15
20
25
30
40
50
60

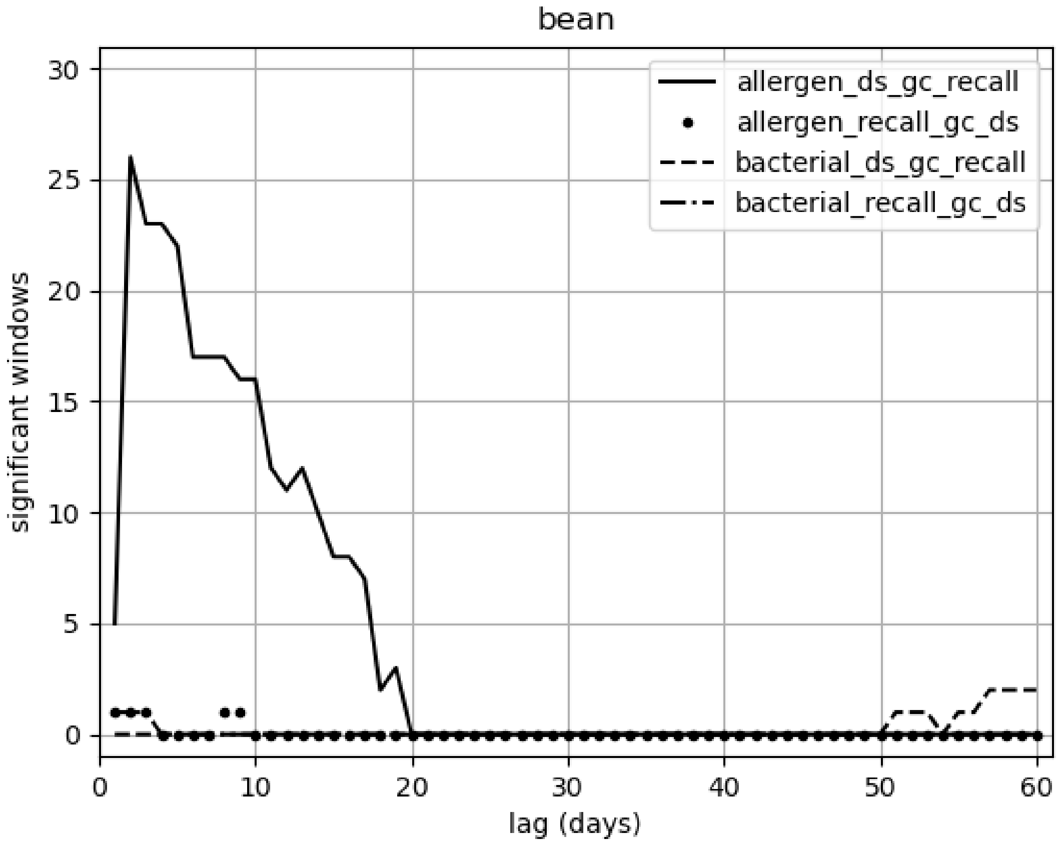
bean
allergen_ds_gc_recall
allergen_recall_gc_ds
bacterial_ds_gc_recall
bacterial_recall_gc_ds
significant windows
lag (days)
0
5
10
15
20
25
30
40
50
60

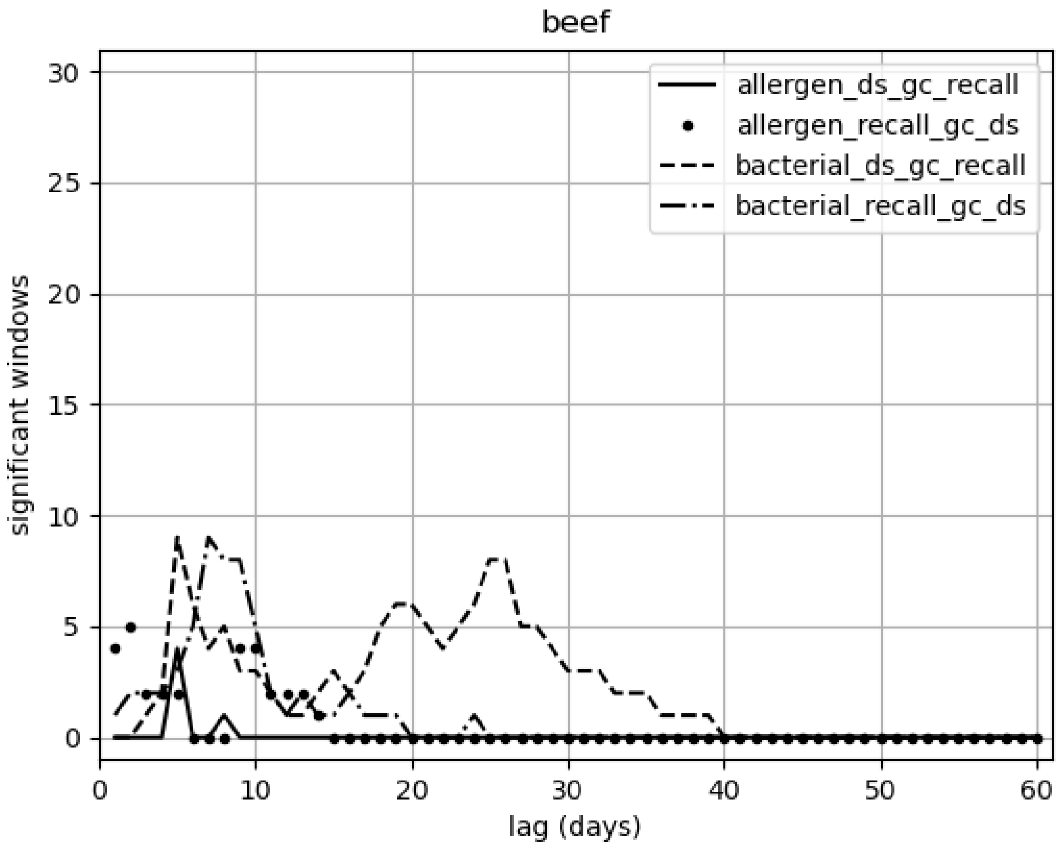

beef
allergen_ds_gc_recall
allergen_recall_gc_ds
bacterial_ds_gc_recall
bacterial_recall_gc_ds
significant windows
lag (days)
0
5
10
15
20
25
30
0
10
20
30
40
50
60

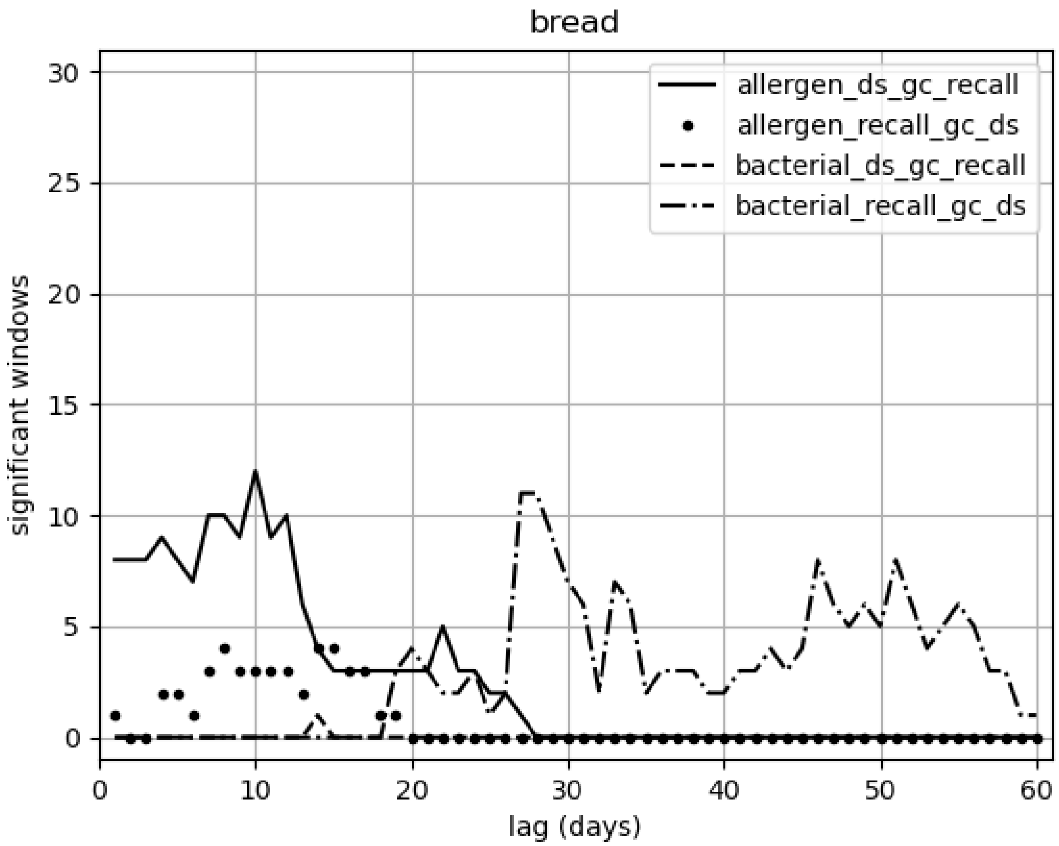

bread
allergen_ds_gc_recall
allergen_recall_gc_ds
bacterial_ds_gc_recall
bacterial_recall_gc_ds
significant windows
lag (days)
0
5
10
15
20
25
30
0
10
20
30
40
50
60

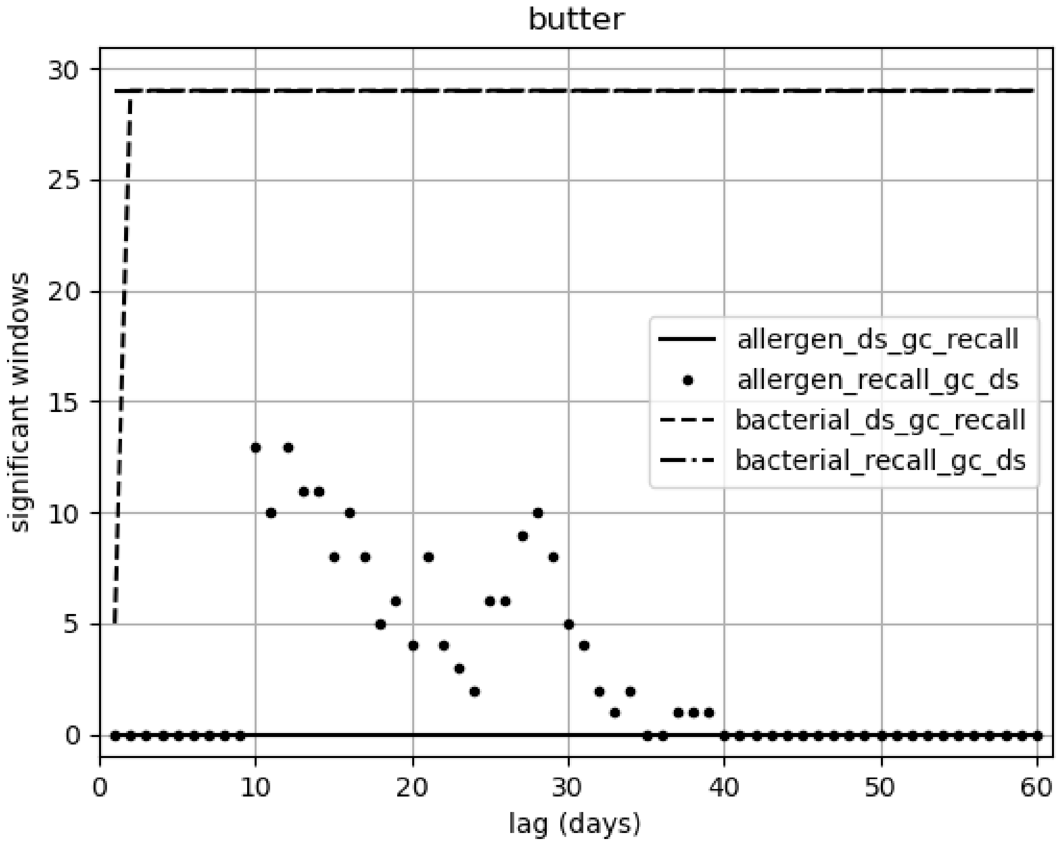
butter
significant windows
lag (days)
allergen_ds_gc_recall
allergen_recall_gc_ds
bacterial_ds_gc_recall
bacterial_recall_gc_ds
0
5
10
15
20
25
30
0
10
20
30
40
50
60

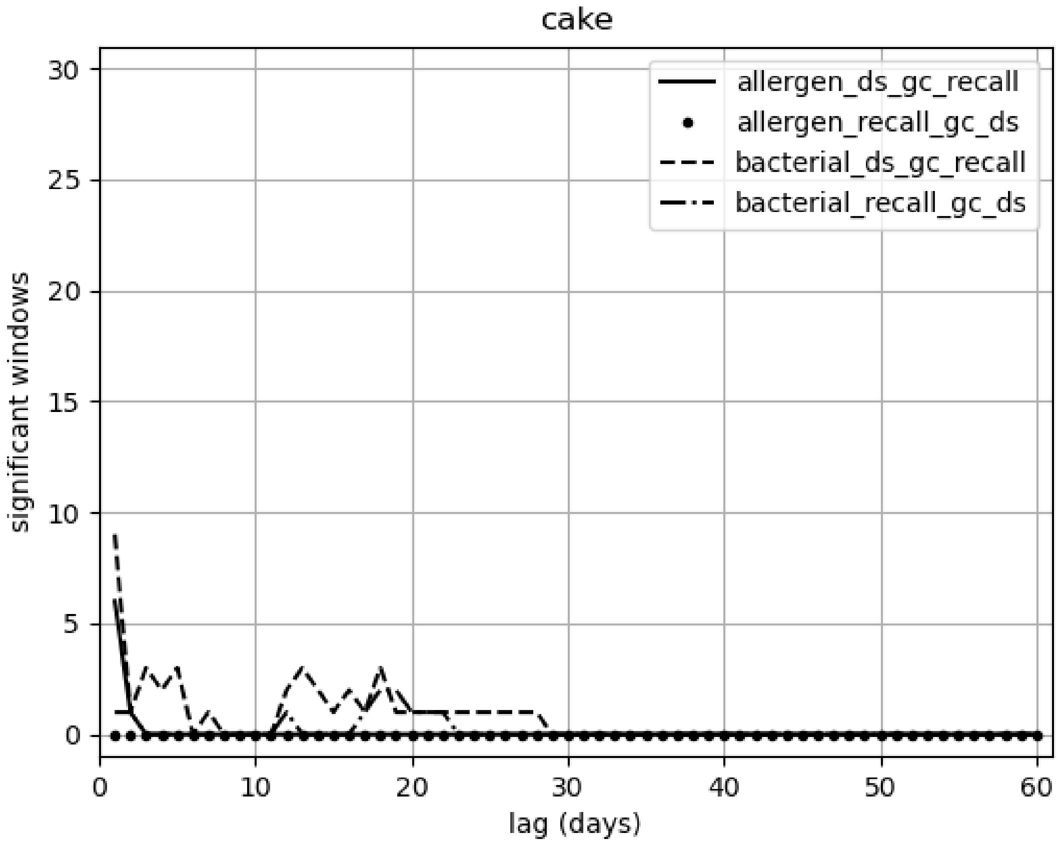
cake
allergen_ds_gc_recall
allergen_recall_gc_ds
bacterial_ds_gc_recall
bacterial_recall_gc_ds
significant windows
lag (days)
0
5
10
15
20
25
30
0
10
20
30
40
50
60

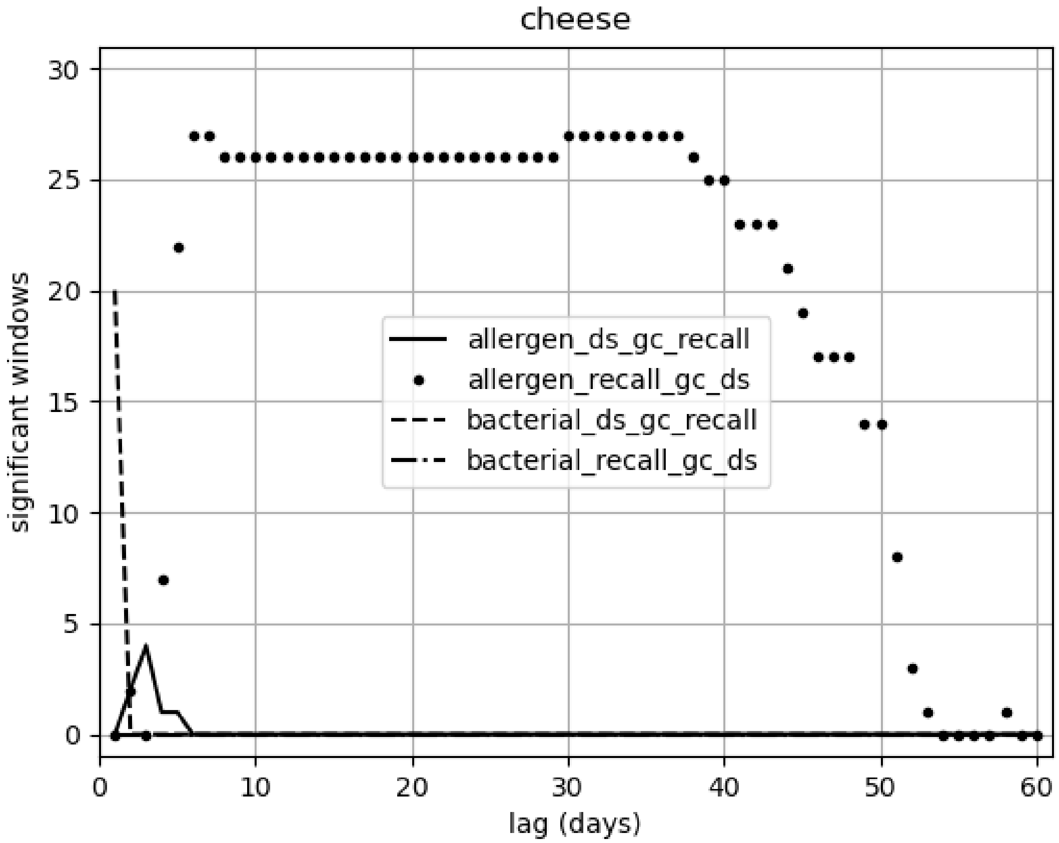

cheese
significant windows
lag (days)
allergen_ds_gc_recall
allergen_recall_gc_ds
bacterial_ds_gc_recall
bacterial_recall_gc_ds
0
5
10
15
20
25
30
0
10
20
30
40
50
60

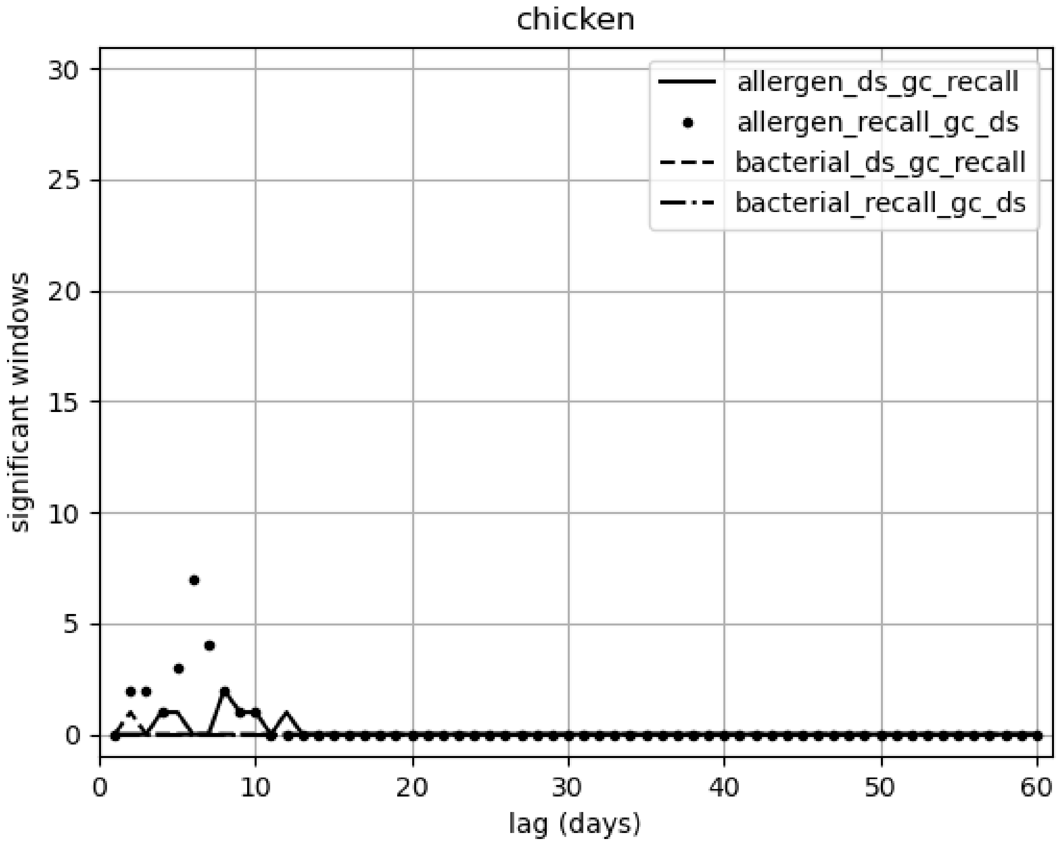
chicken
allergen_ds_gc_recall
allergen_recall_gc_ds
bacterial_ds_gc_recall
bacterial_recall_gc_ds
significant windows
lag (days)
0
5
10
15
20
25
30
40
50
60

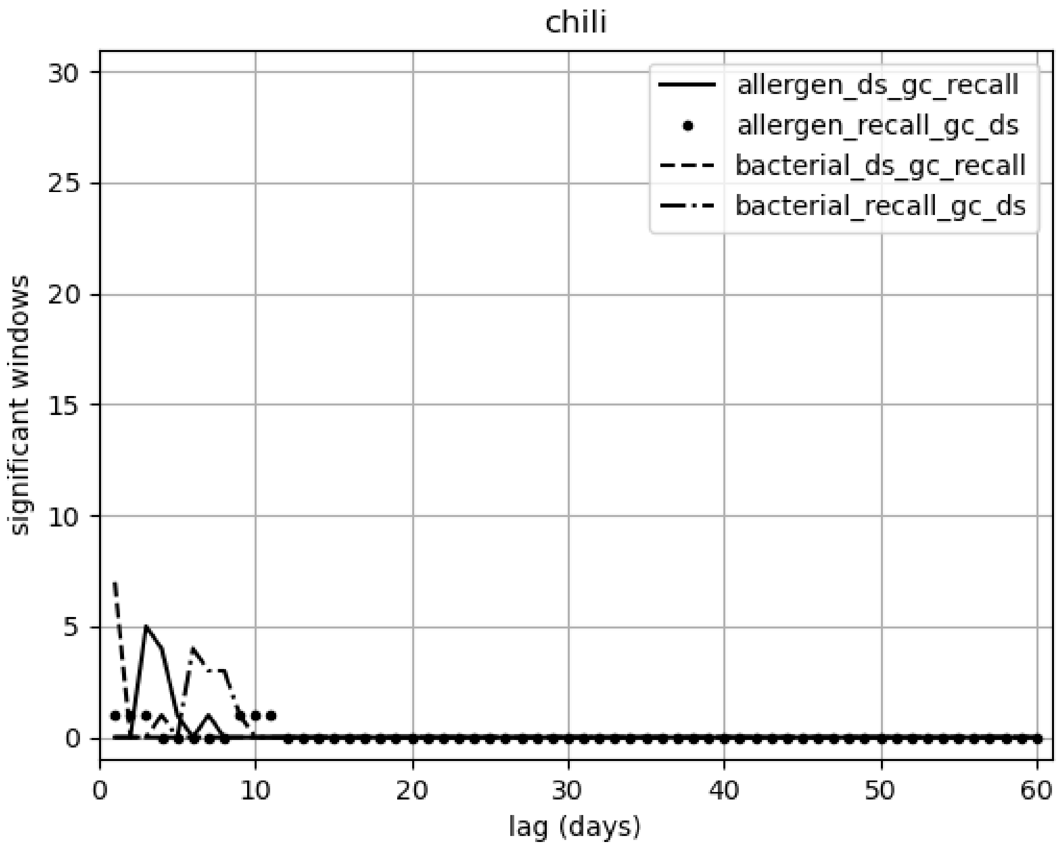
chili
allergen_ds_gc_recall
allergen_recall_gc_ds
bacterial_ds_gc_recall
bacterial_recall_gc_ds
significant windows
lag (days)
0
5
10
15
20
25
30
40
50
60

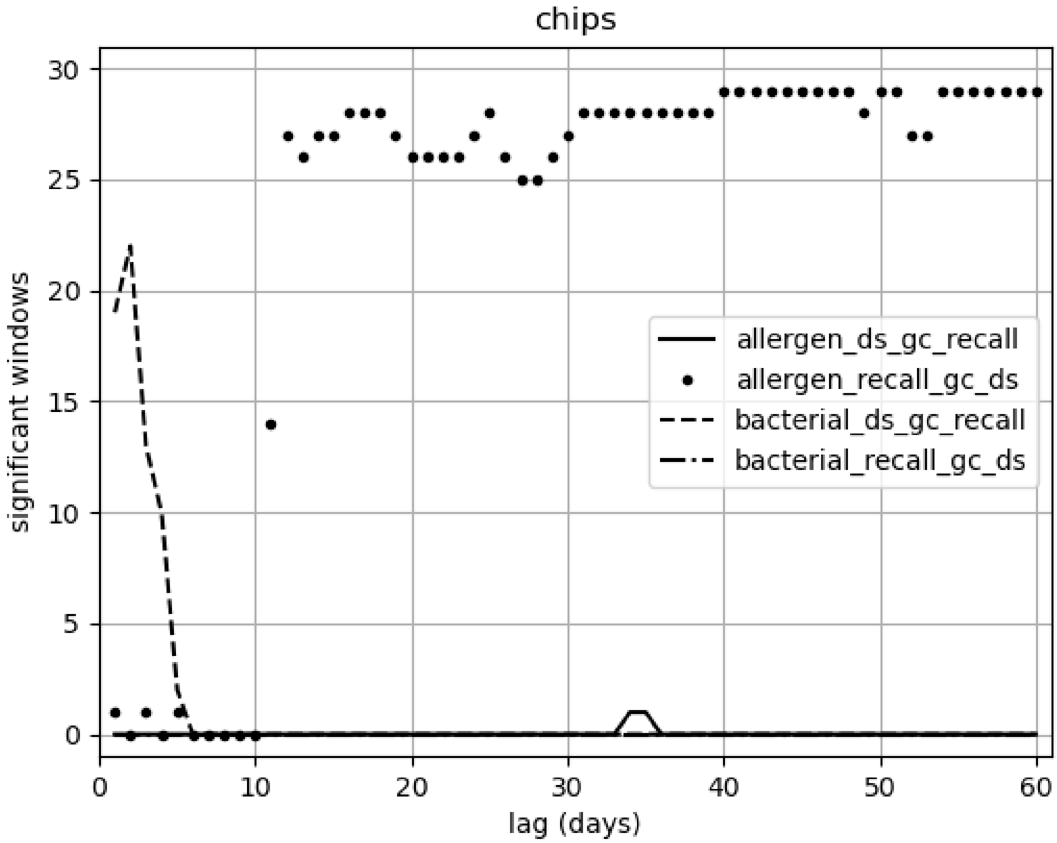
chips
significant windows
lag (days)
allergen_ds_gc_recall
allergen_recall_gc_ds
bacterial_ds_gc_recall
bacterial_recall_gc_ds
0
5
10
15
20
25
30
0
10
20
30
40
50
60

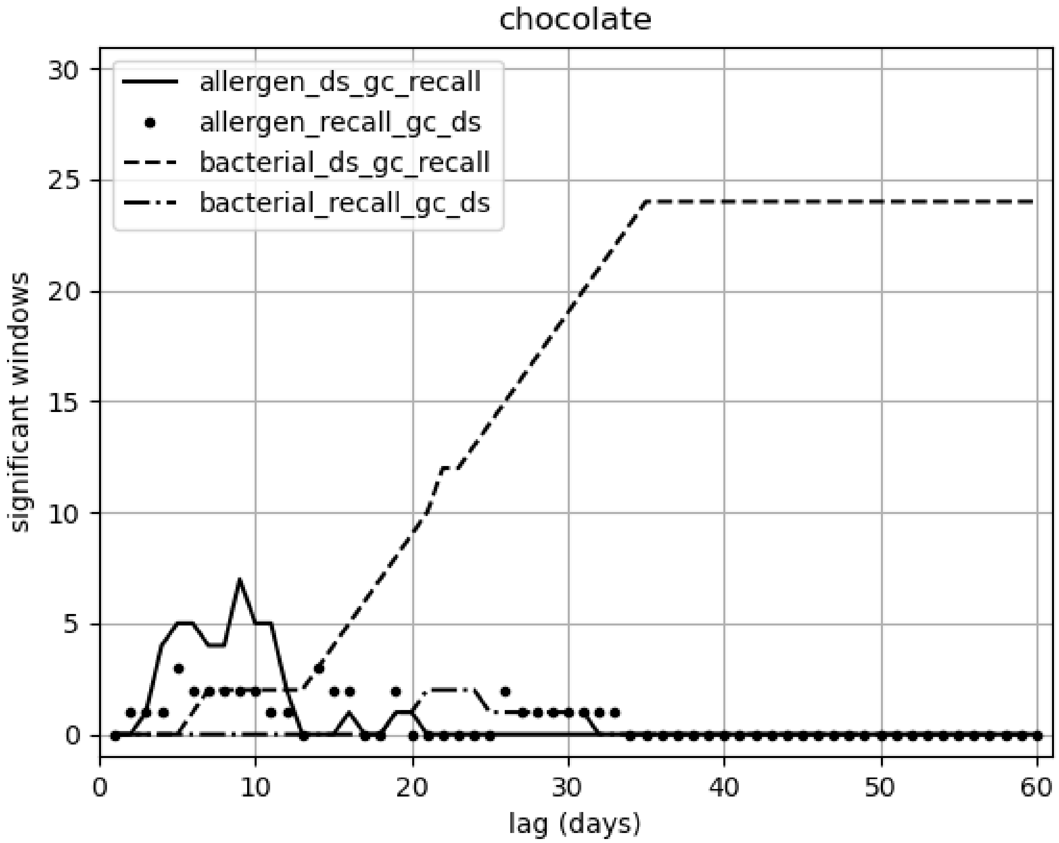

chocolate
allergen_ds_gc_recall
allergen_recall_gc_ds
bacterial_ds_gc_recall
bacterial_recall_gc_ds
significant windows
lag (days)
0
5
10
15
20
25
30
0
10
20
30
40
50
60

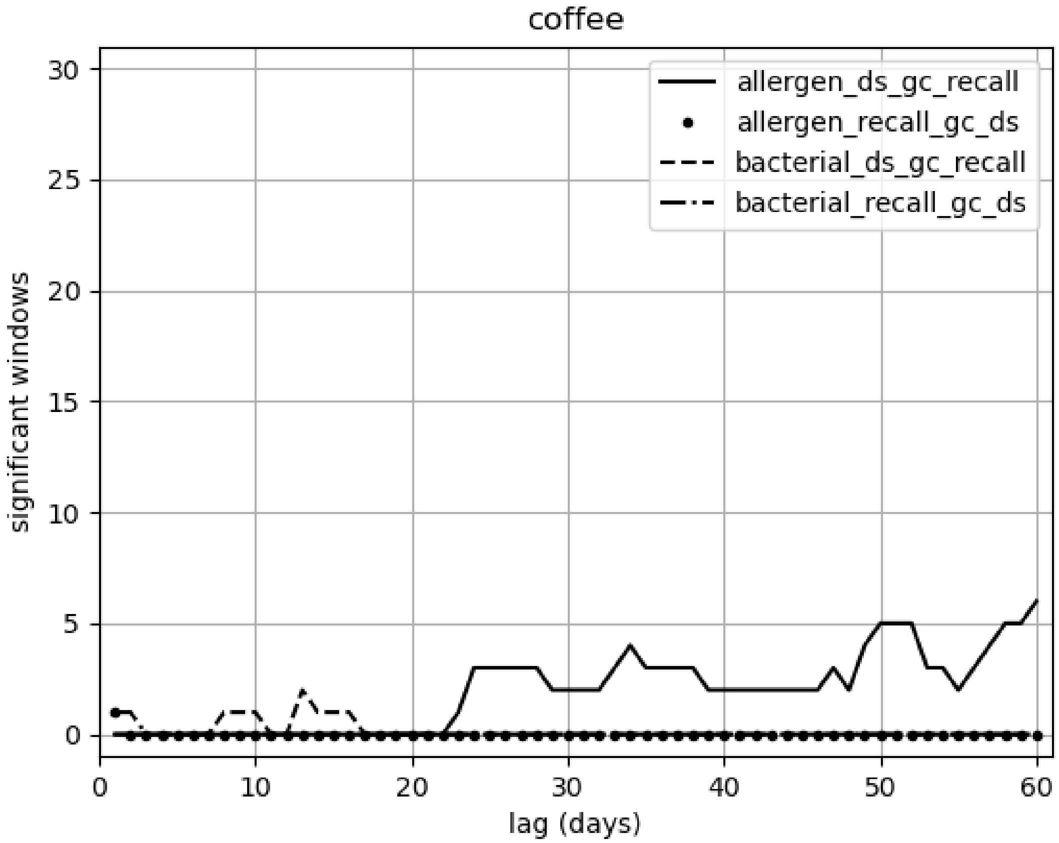

coffee
allergen_ds_gc_recall
allergen_recall_gc_ds
bacterial_ds_gc_recall
bacterial_recall_gc_ds
significant windows
lag (days)
0
5
10
15
20
25
30
0
10
20
30
40
50
60

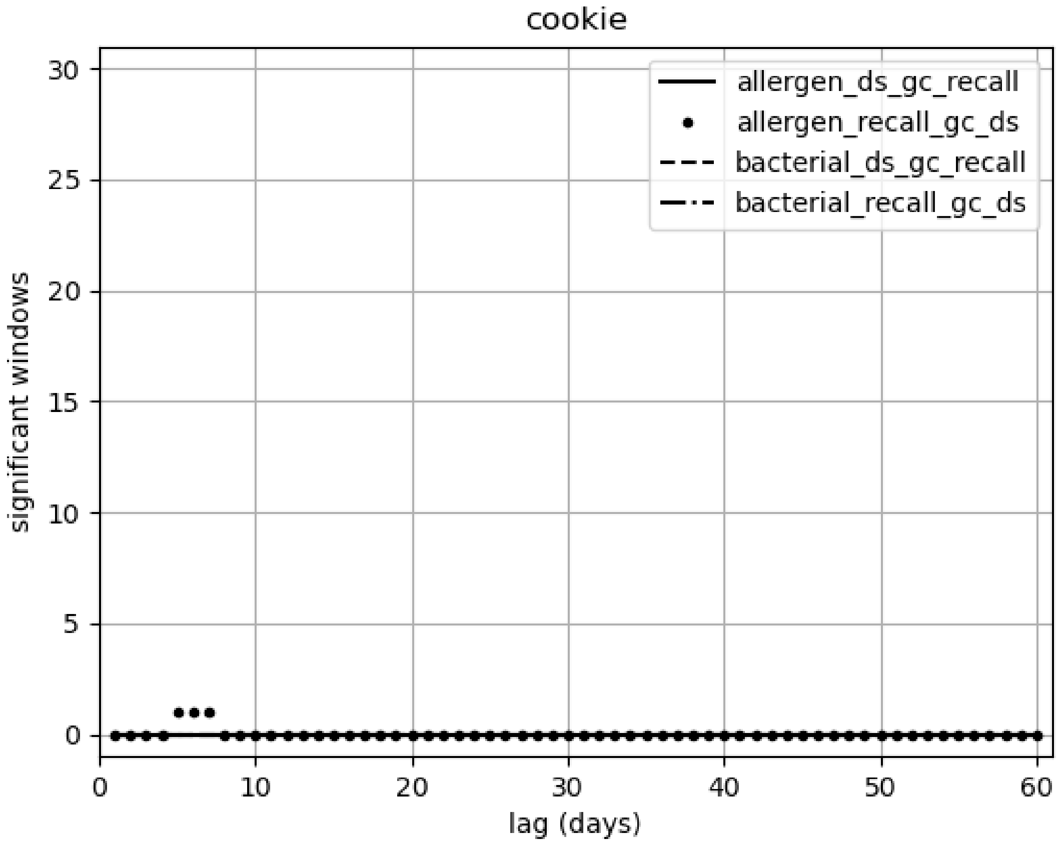

cookie
allergen_ds_gc_recall
allergen_recall_gc_ds
bacterial_ds_gc_recall
bacterial_recall_gc_ds
significant windows
lag (days)
0
5
10
15
20
25
30
40
50
60

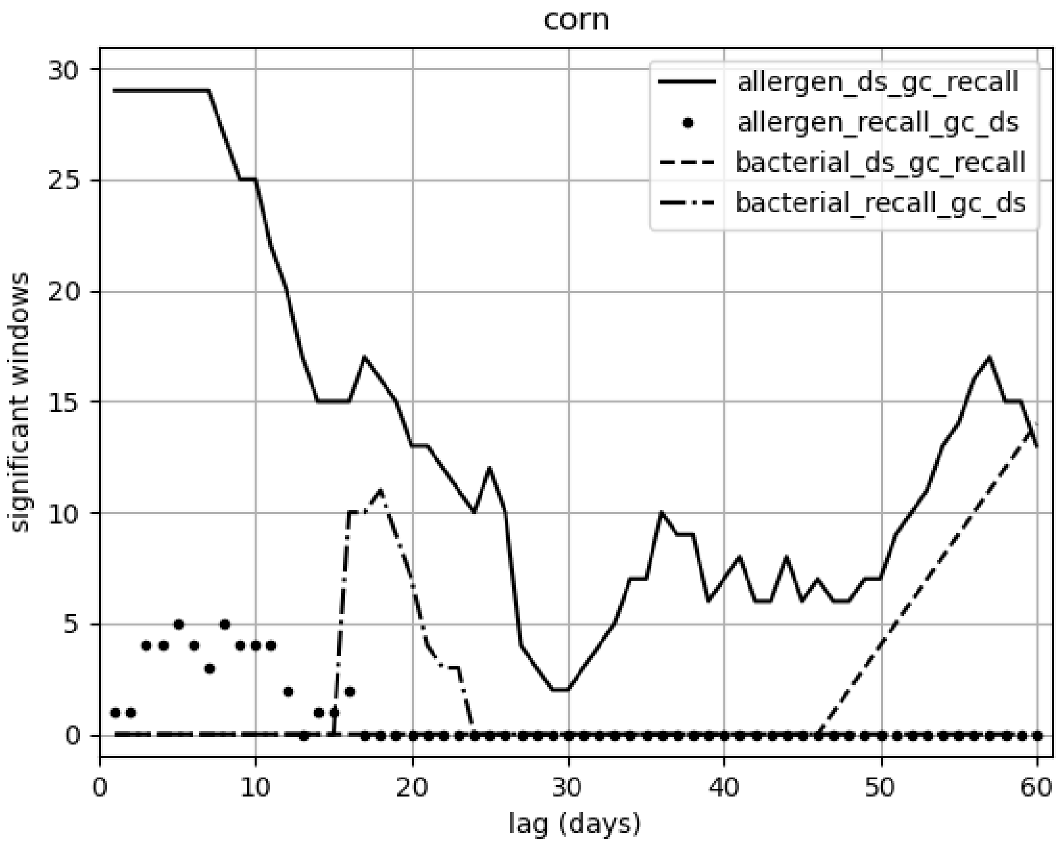

corn
allergen_ds_gc_recall
allergen_recall_gc_ds
bacterial_ds_gc_recall
bacterial_recall_gc_ds
significant windows
lag (days)
0
5
10
15
20
25
30
40
50
60

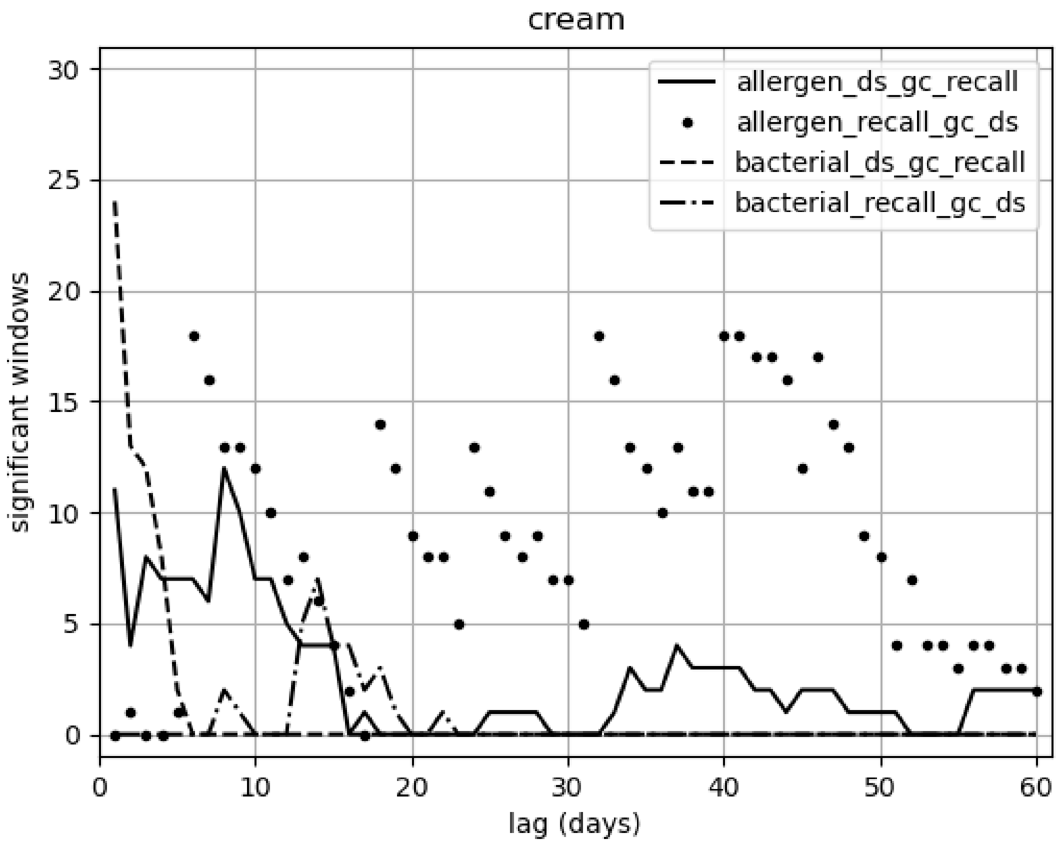
cream
allergen_ds_gc_recall
allergen_recall_gc_ds
bacterial_ds_gc_recall
bacterial_recall_gc_ds
significant windows
lag (days)
0
5
10
15
20
25
30
0
10
20
30
40
50
60

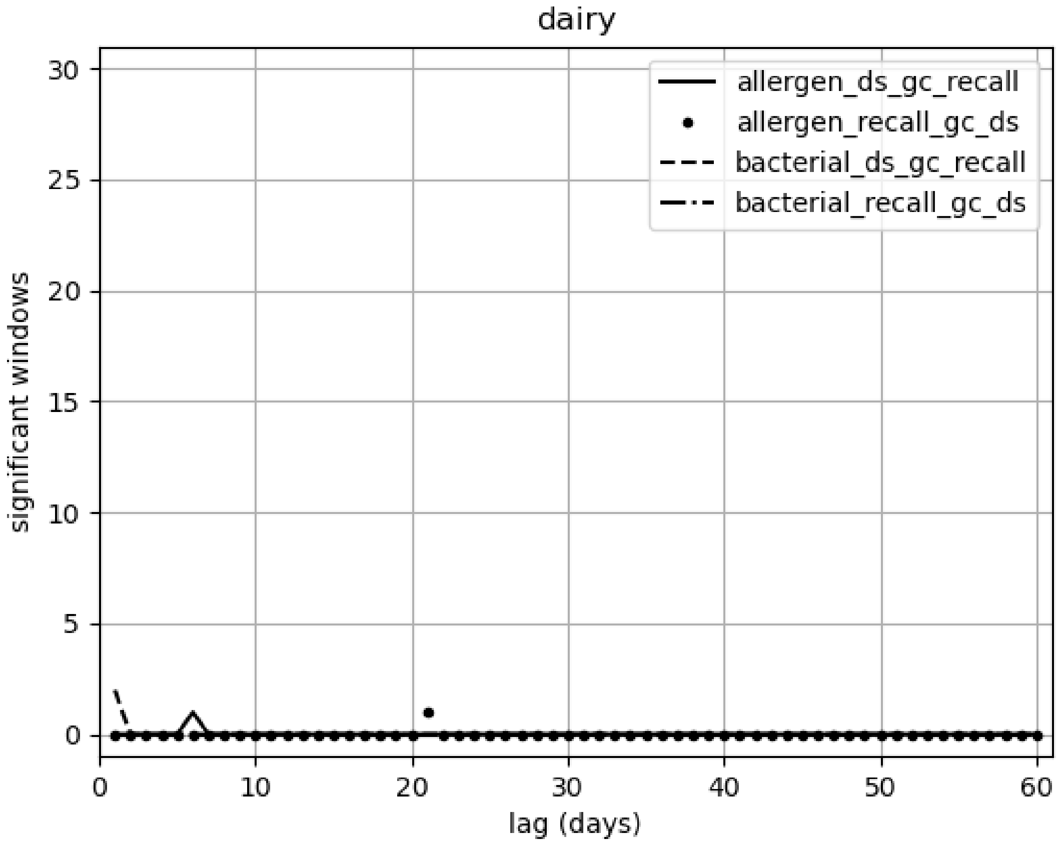
dairy
allergen_ds_gc_recall
allergen_recall_gc_ds
bacterial_ds_gc_recall
bacterial_recall_gc_ds
significant windows
lag (days)
0
5
10
15
20
25
30
40
50
60

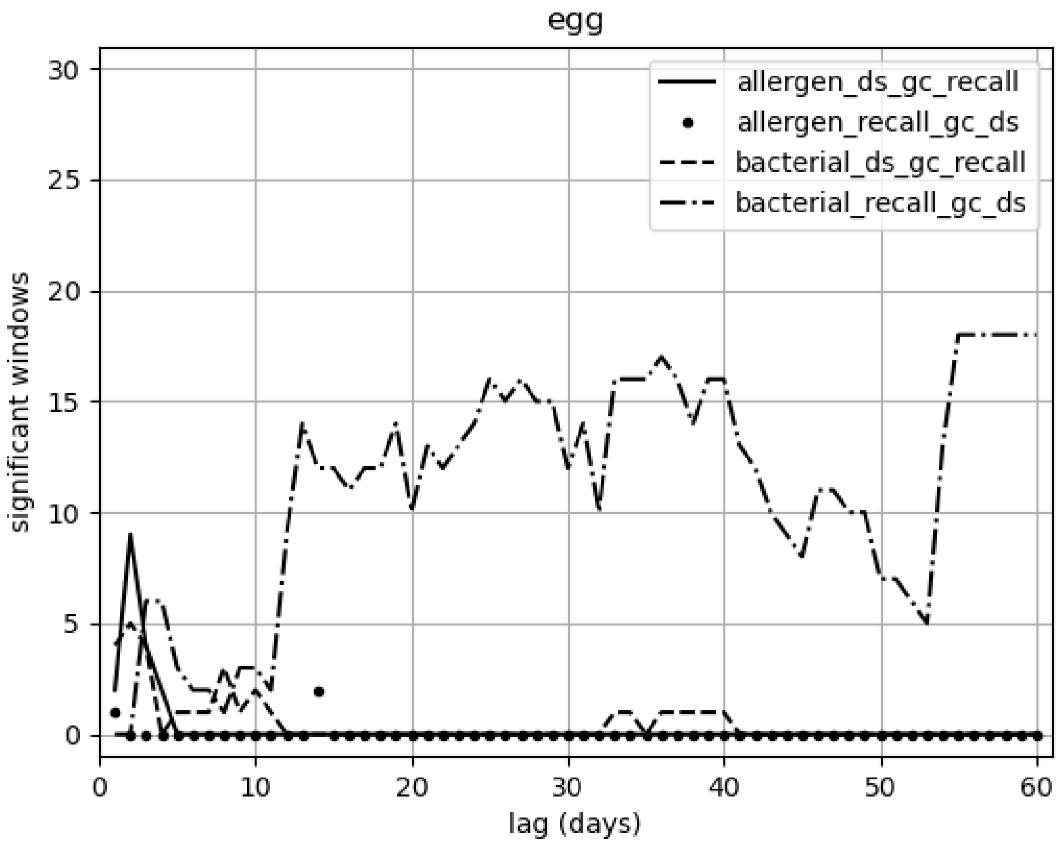

egg
allergen_ds_gc_recall
allergen_recall_gc_ds
bacterial_ds_gc_recall
bacterial_recall_gc_ds
significant windows
lag (days)
0
5
10
15
20
25
30
0
10
20
30
40
50
60

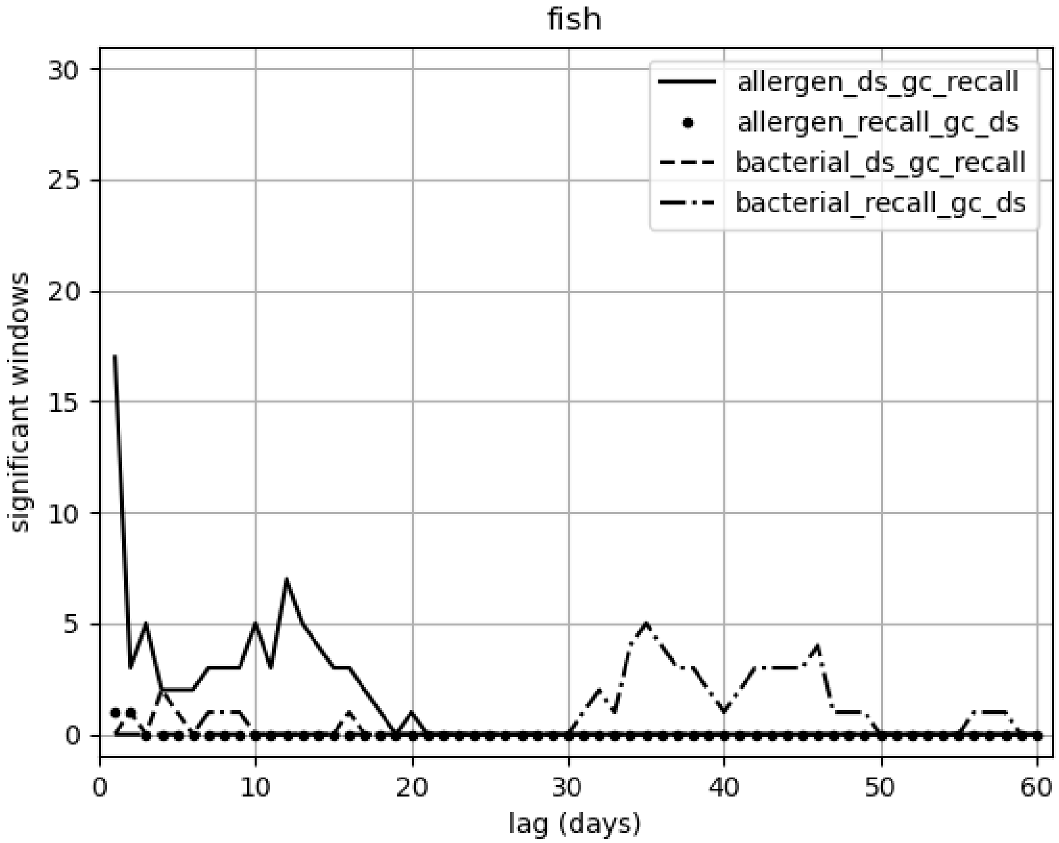

fish
allergen_ds_gc_recall
allergen_recall_gc_ds
bacterial_ds_gc_recall
bacterial_recall_gc_ds
significant windows
lag (days)
0
5
10
15
20
25
30
40
50
60

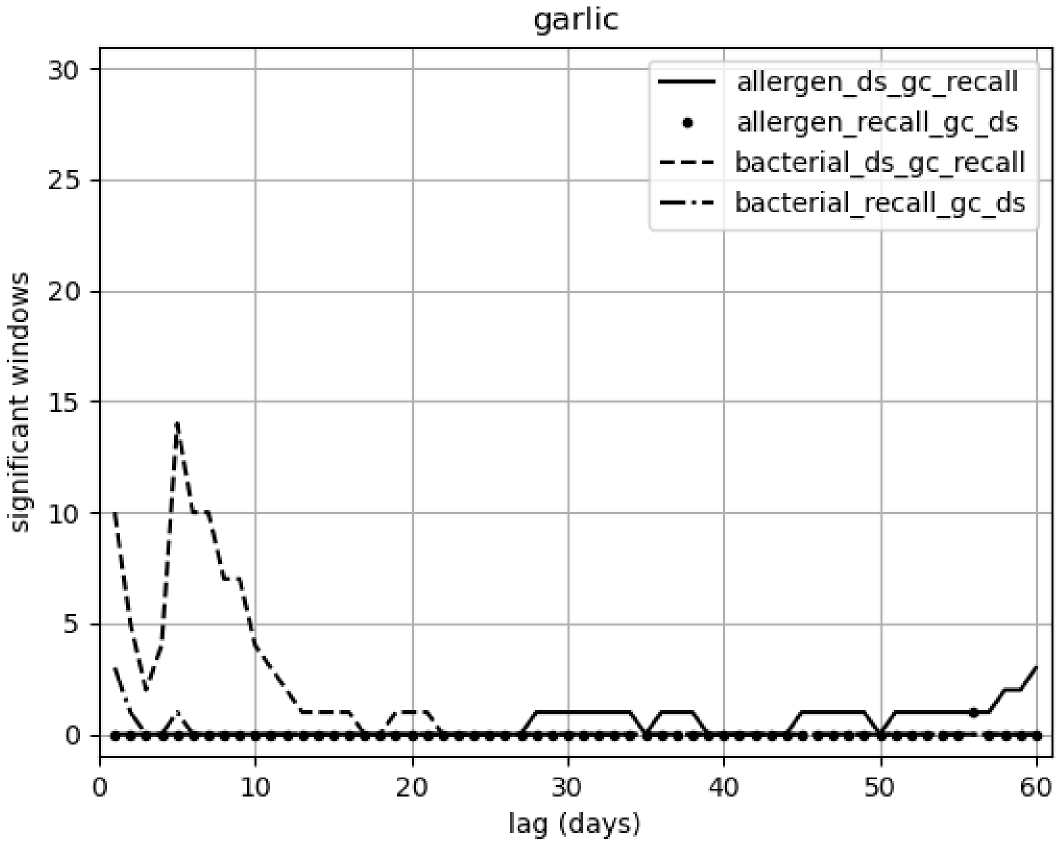
garlic
allergen_ds_gc_recall
allergen_recall_gc_ds
bacterial_ds_gc_recall
bacterial_recall_gc_ds
significant windows
lag (days)
0
5
10
15
20
25
30
40
50
60

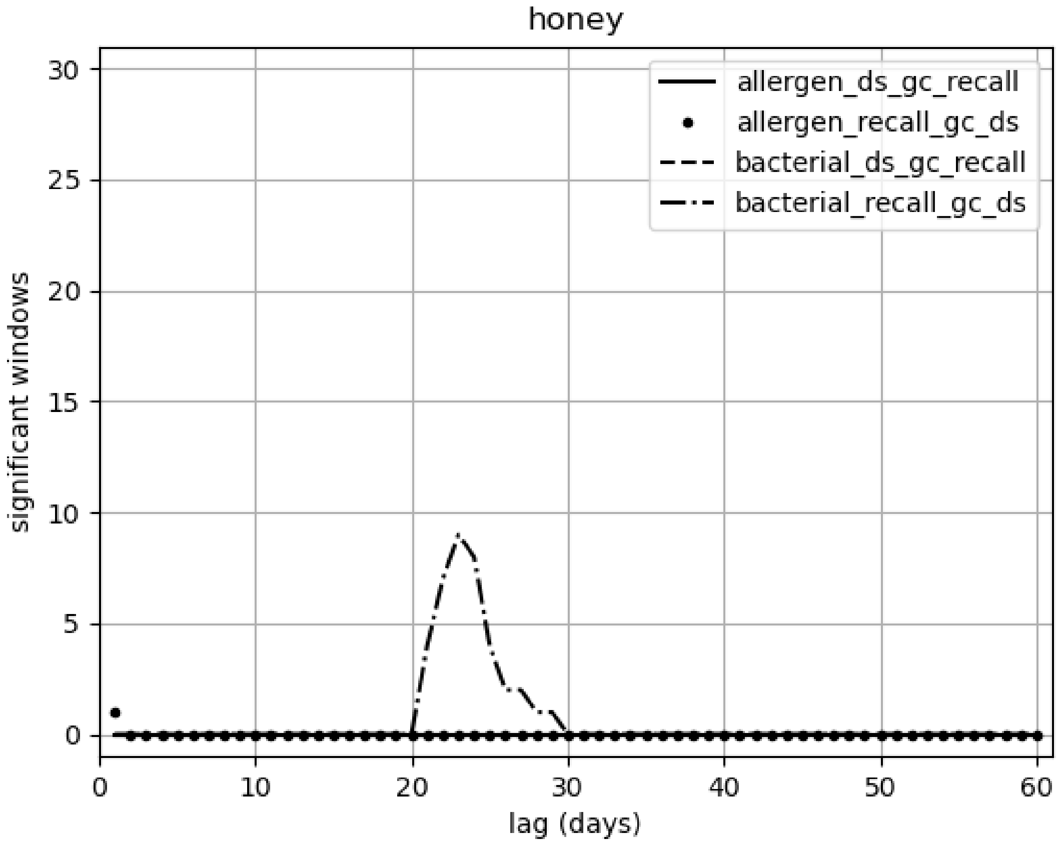

honey
allergen_ds_gc_recall
allergen_recall_gc_ds
bacterial_ds_gc_recall
bacterial_recall_gc_ds
significant windows
lag (days)
0
5
10
15
20
25
30
0
10
20
30
40
50
60

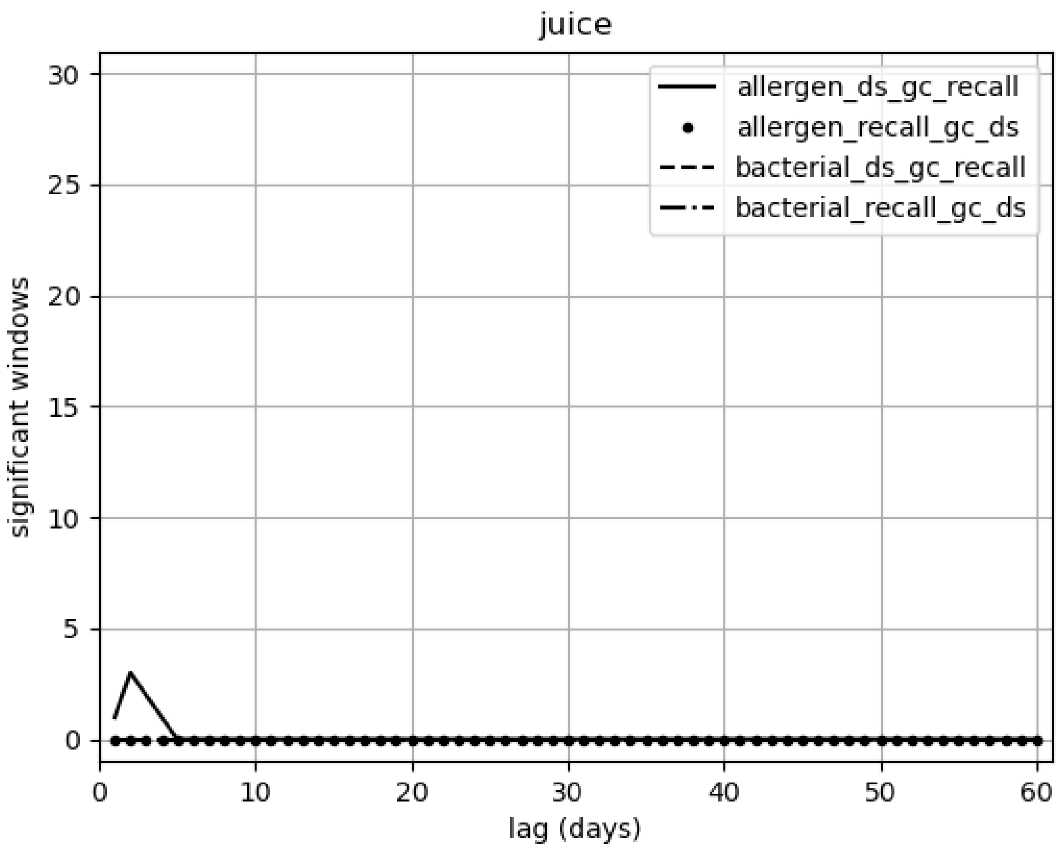
juice
allergen_ds_gc_recall
allergen_recall_gc_ds
bacterial_ds_gc_recall
bacterial_recall_gc_ds
significant windows
lag (days)
0
5
10
15
20
25
30
0
10
20
30
40
50
60

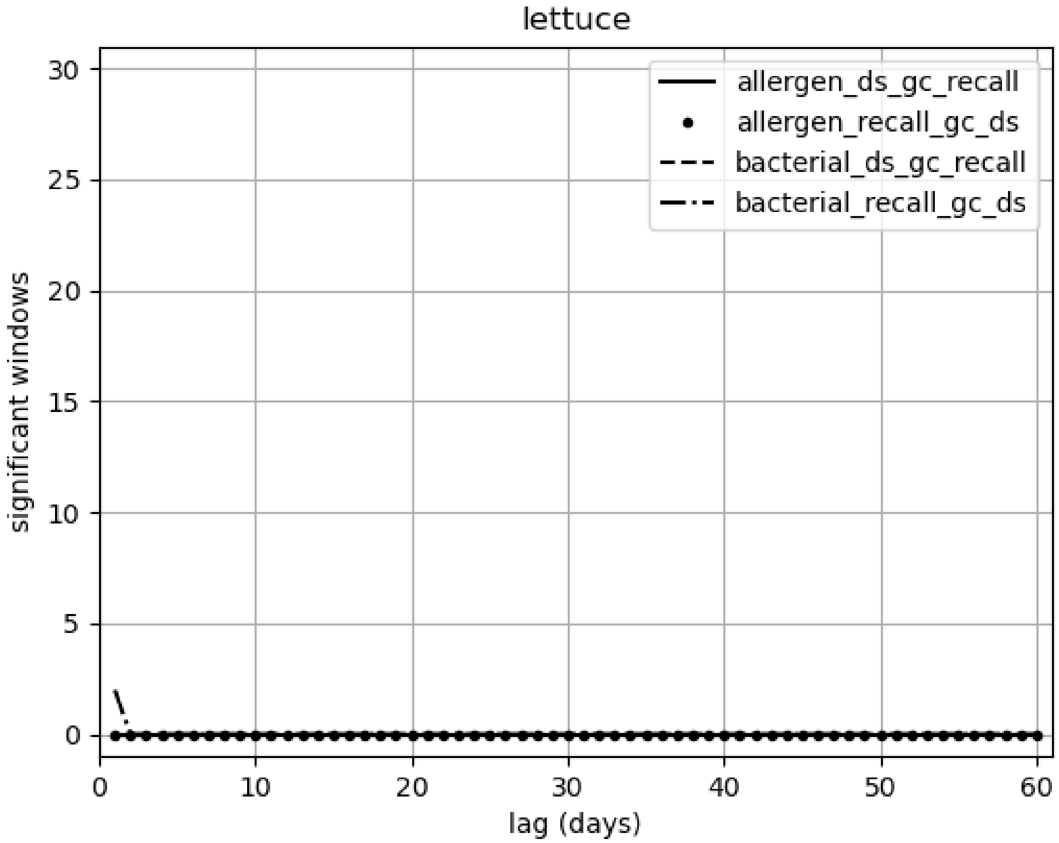

lettuce
allergen_ds_gc_recall
allergen_recall_gc_ds
bacterial_ds_gc_recall
bacterial_recall_gc_ds
significant windows
lag (days)
0
5
10
15
20
25
30
0
10
20
30
40
50
60

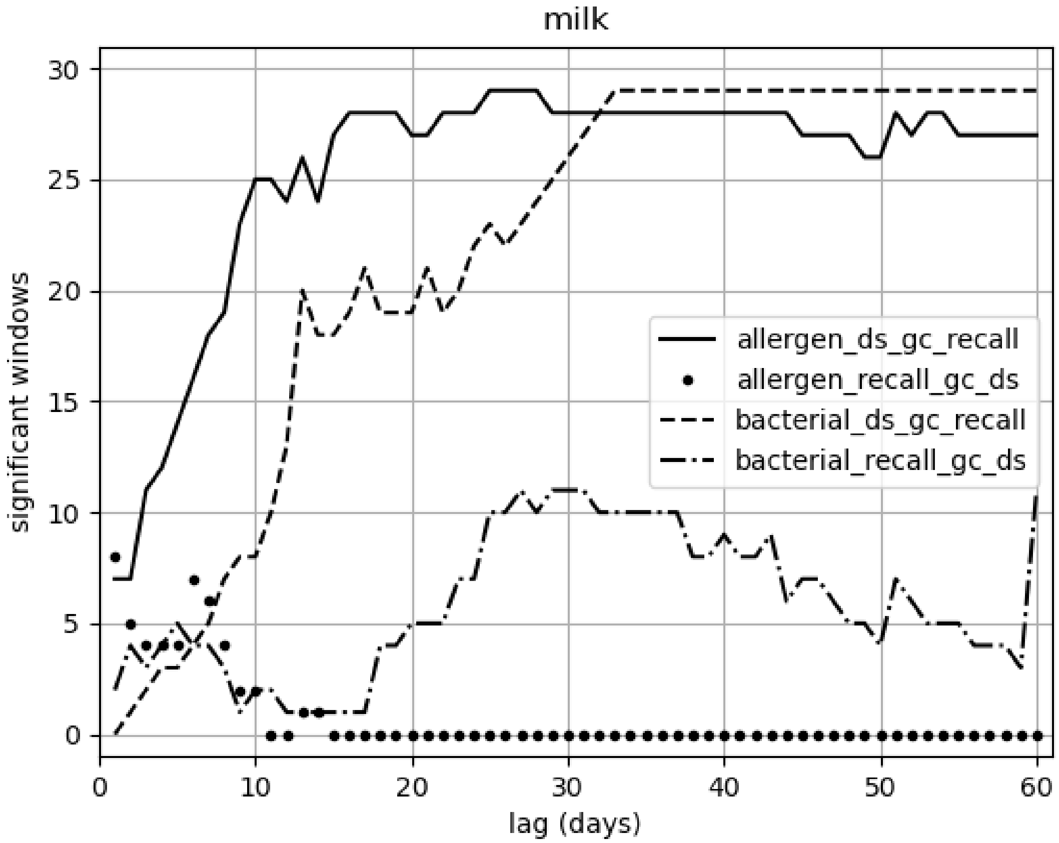
milk
significant windows
lag (days)
allergen_ds_gc_recall
allergen_recall_gc_ds
bacterial_ds_gc_recall
bacterial_recall_gc_ds
0
5
10
15
20
25
30
40
50
60

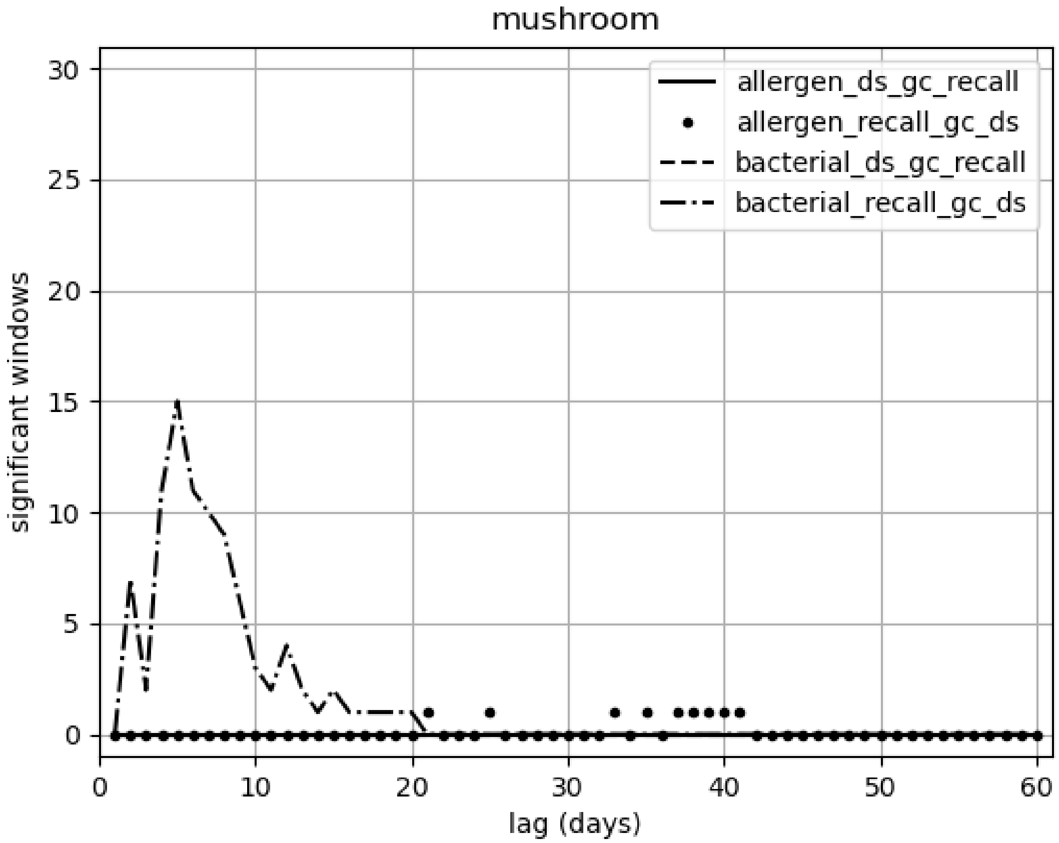
mushroom
allergen_ds_gc_recall
allergen_recall_gc_ds
bacterial_ds_gc_recall
bacterial_recall_gc_ds
significant windows
lag (days)
0
5
10
15
20
25
30
0
10
20
30
40
50
60

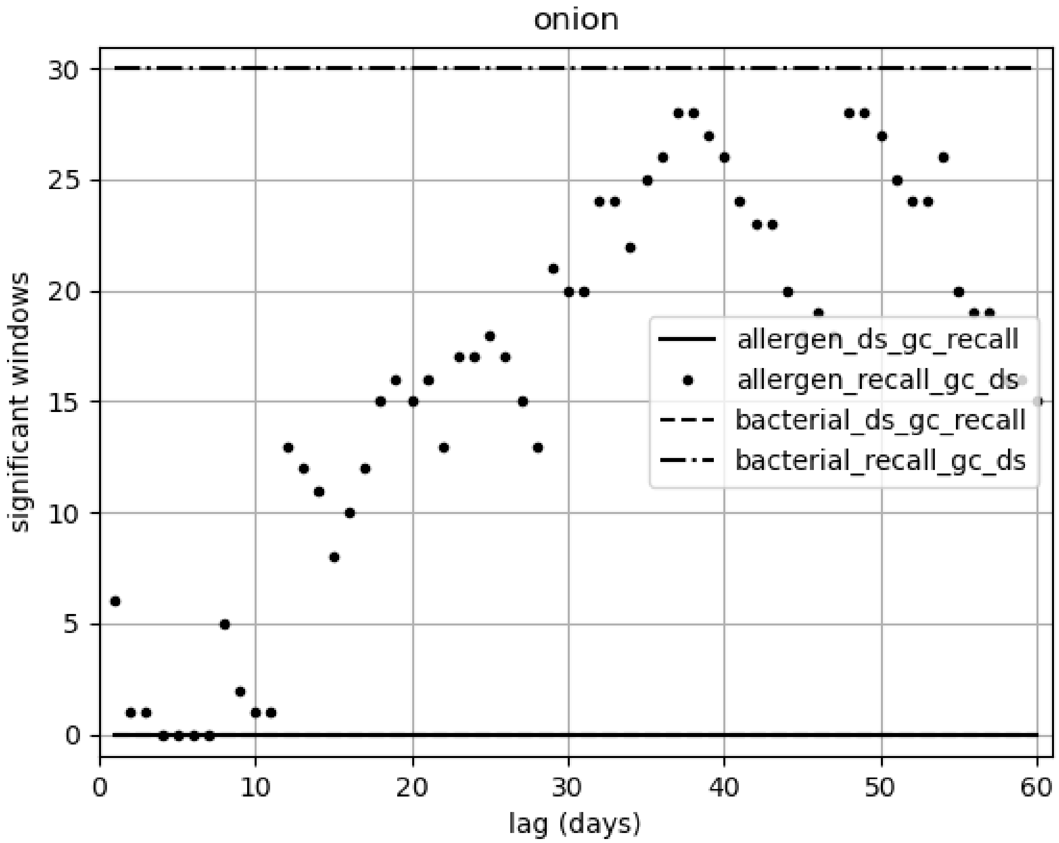

onion
significant windows
lag (days)
allergen_ds_gc_recall
allergen_recall_gc_ds
bacterial_ds_gc_recall
bacterial_recall_gc_ds
0
5
10
15
20
25
30
0
10
20
30
40
50
60

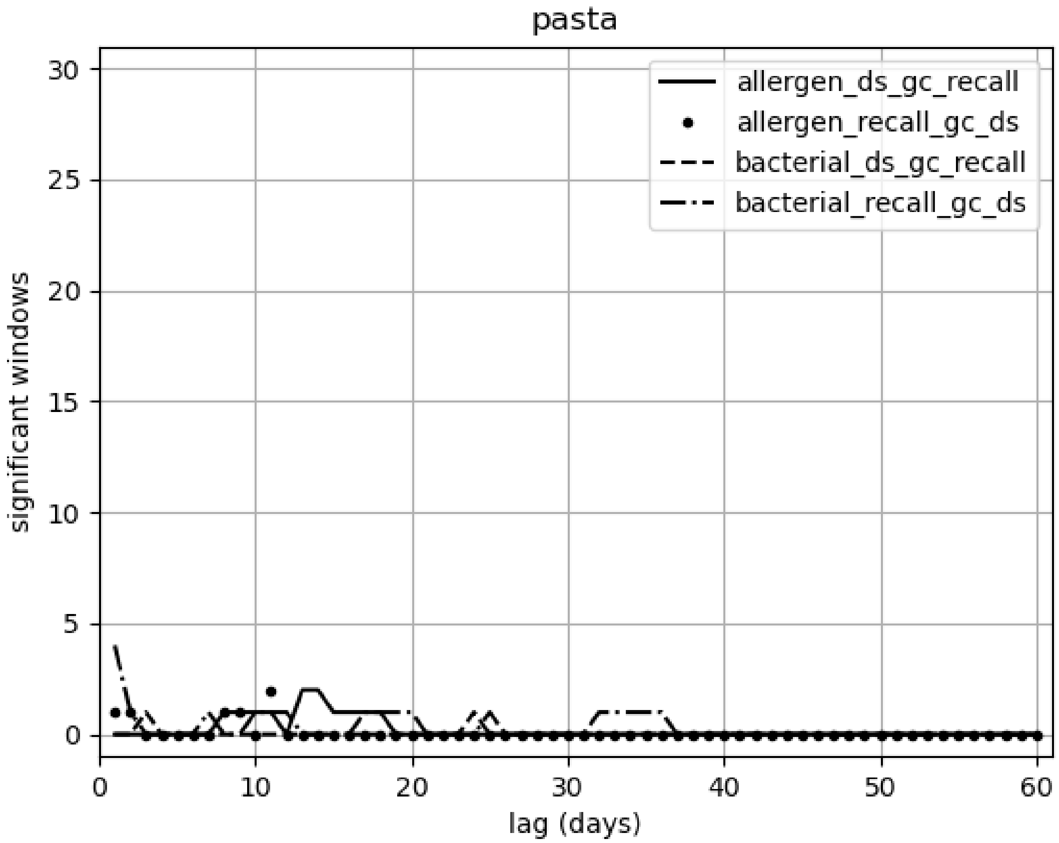
pasta
allergen_ds_gc_recall
allergen_recall_gc_ds
bacterial_ds_gc_recall
bacterial_recall_gc_ds
significant windows
lag (days)
0
5
10
15
20
25
30
0
10
20
30
40
50
60

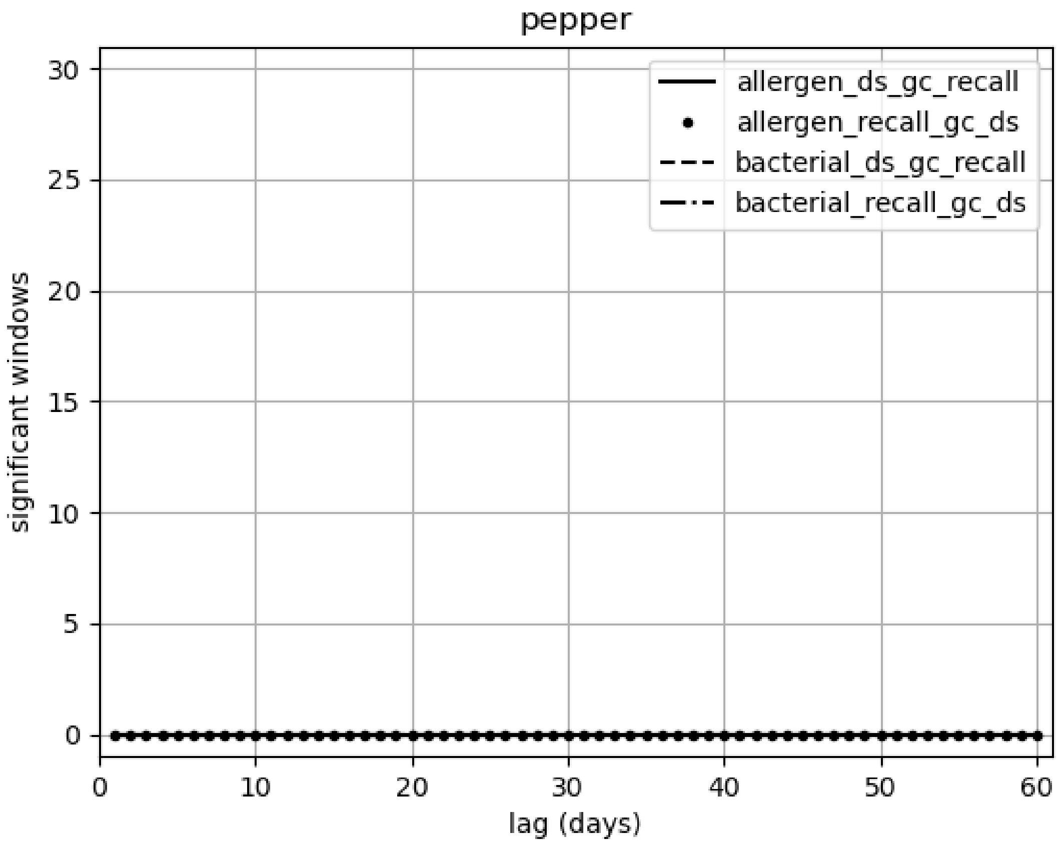
pepper
allergen_ds_gc_recall
allergen_recall_gc_ds
bacterial_ds_gc_recall
bacterial_recall_gc_ds
significant windows
lag (days)
0
5
10
15
20
25
30
0
10
20
30
40
50
60

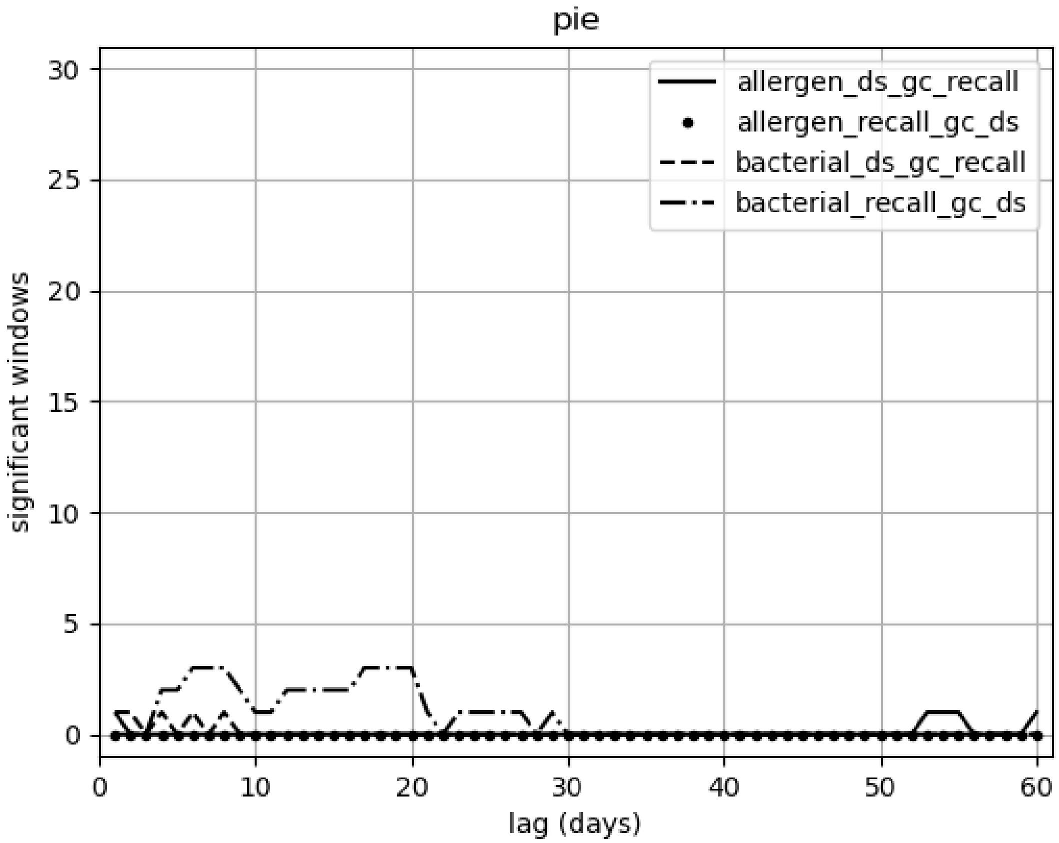
pie
allergen_ds_gc_recall
allergen_recall_gc_ds
bacterial_ds_gc_recall
bacterial_recall_gc_ds
significant windows
lag (days)
0
5
10
15
20
25
30
40
50
60

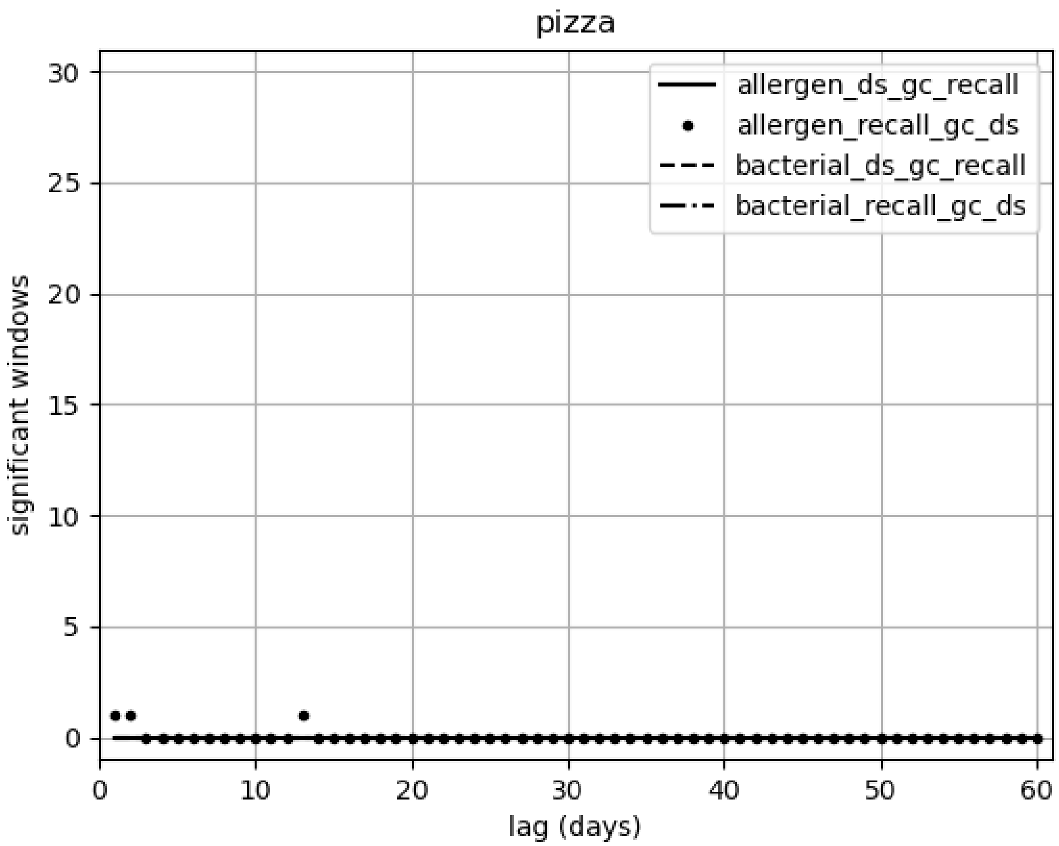

pizza
allergen_ds_gc_recall
allergen_recall_gc_ds
bacterial_ds_gc_recall
bacterial_recall_gc_ds
significant windows
lag (days)
0
5
10
15
20
25
30
40
50
60

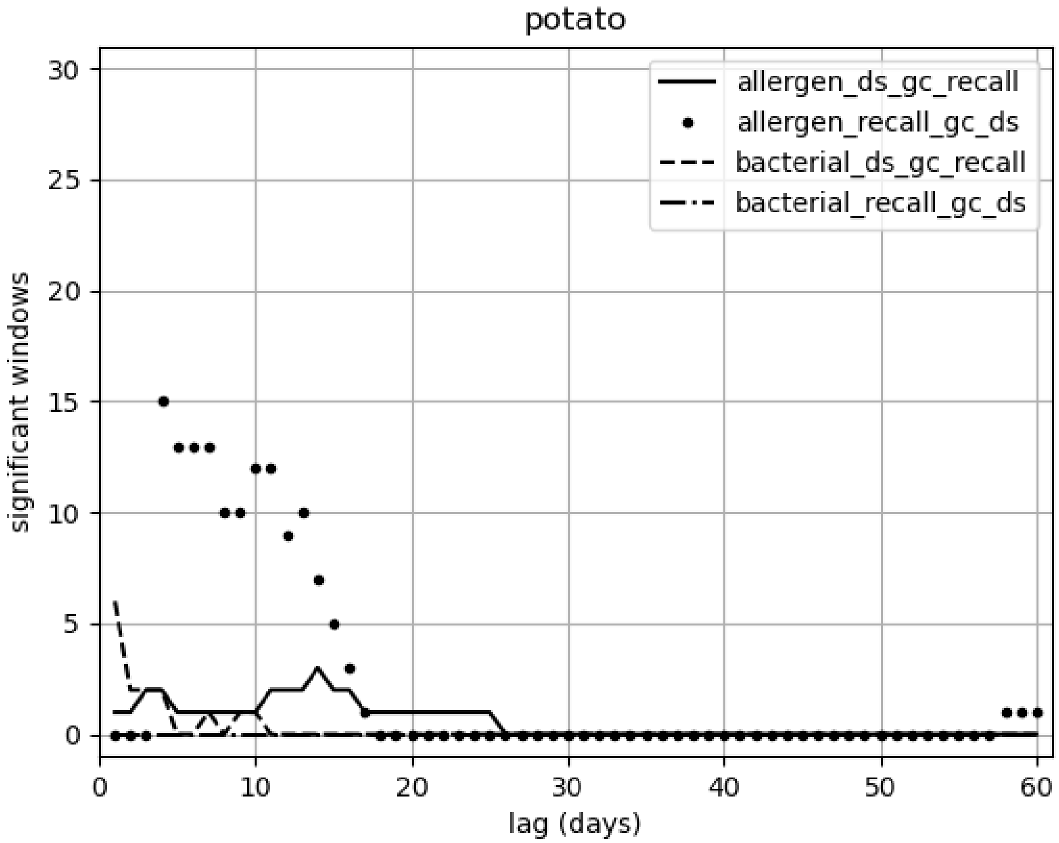

potato
allergen_ds_gc_recall
allergen_recall_gc_ds
bacterial_ds_gc_recall
bacterial_recall_gc_ds
significant windows
lag (days)
0
5
10
15
20
25
30
40
50
60

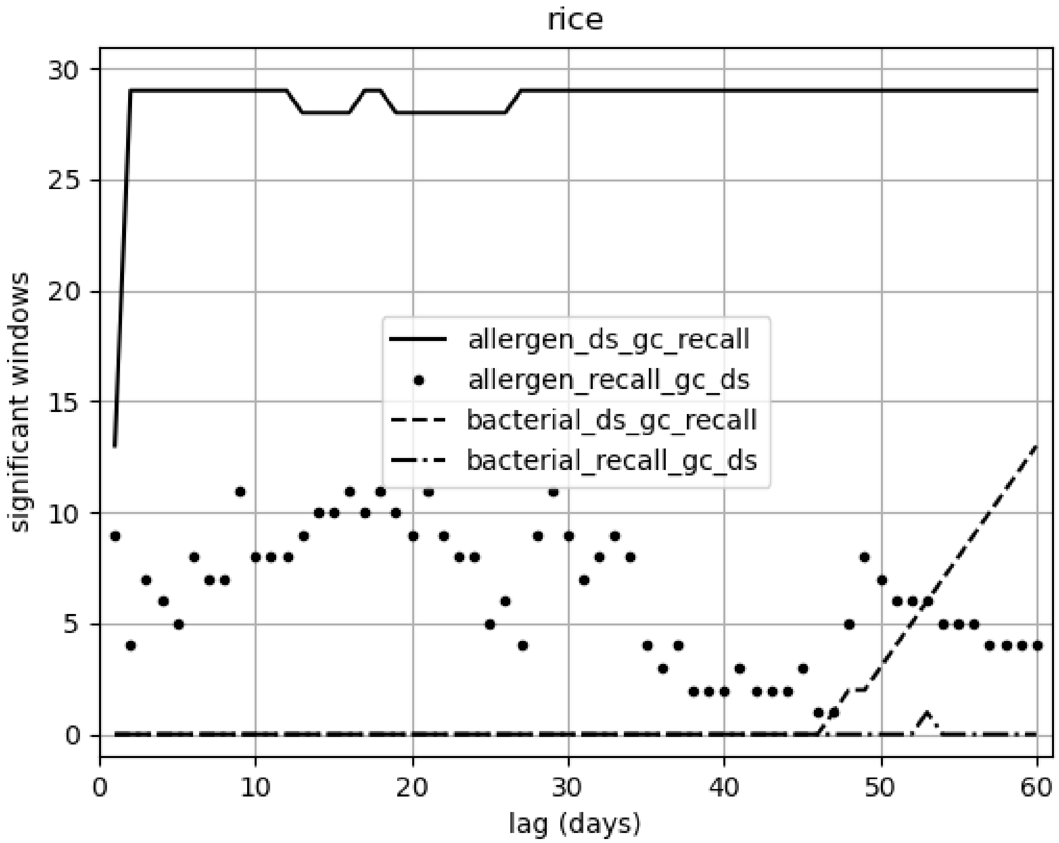
rice
significant windows
lag (days)
allergen_ds_gc_recall
allergen_recall_gc_ds
bacterial_ds_gc_recall
bacterial_recall_gc_ds

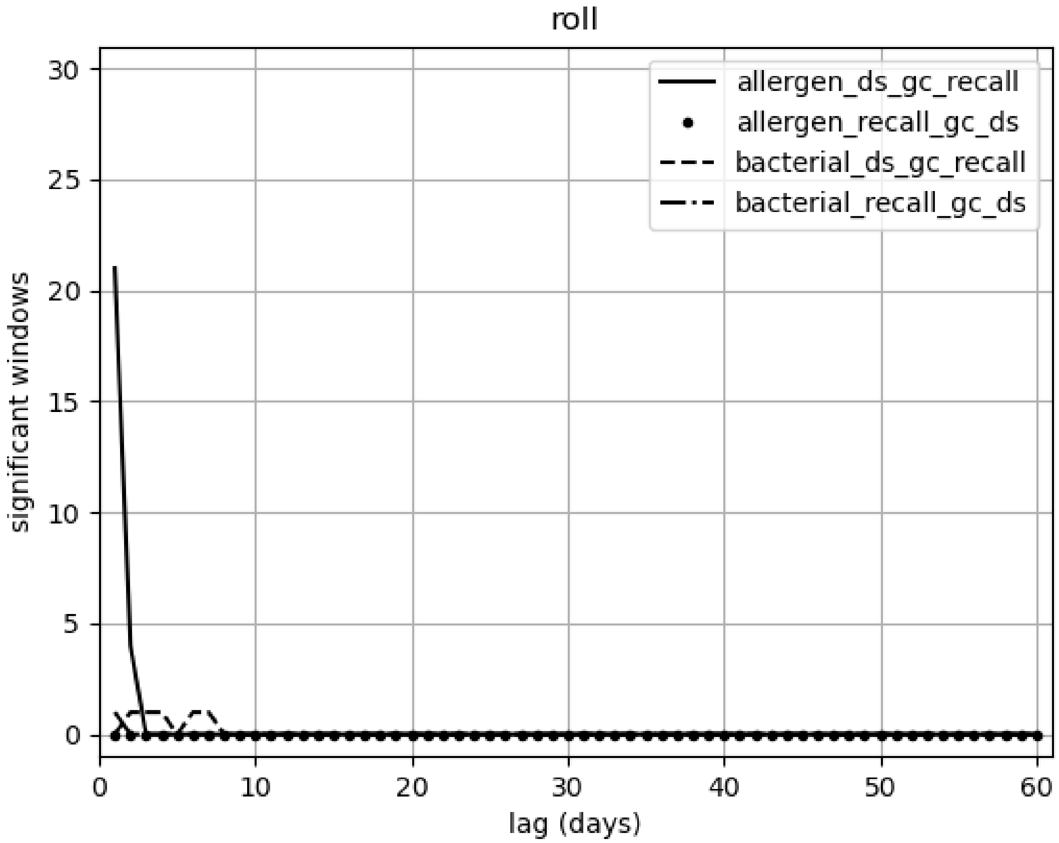

roll
allergen_ds_gc_recall
allergen_recall_gc_ds
bacterial_ds_gc_recall
bacterial_recall_gc_ds
significant windows
lag (days)
0
5
10
15
20
25
30
0
10
20
30
40
50
60

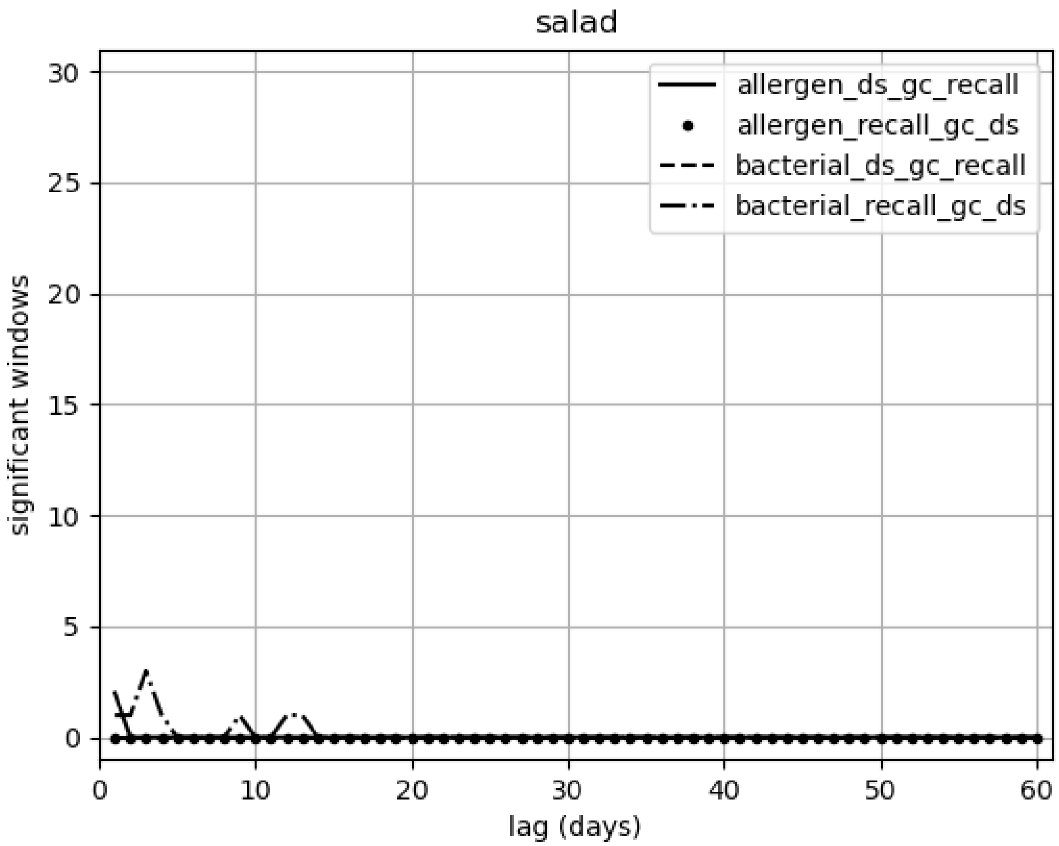

salad
allergen_ds_gc_recall
allergen_recall_gc_ds
bacterial_ds_gc_recall
bacterial_recall_gc_ds
significant windows
lag (days)
0
5
10
15
20
25
30
0
10
20
30
40
50
60

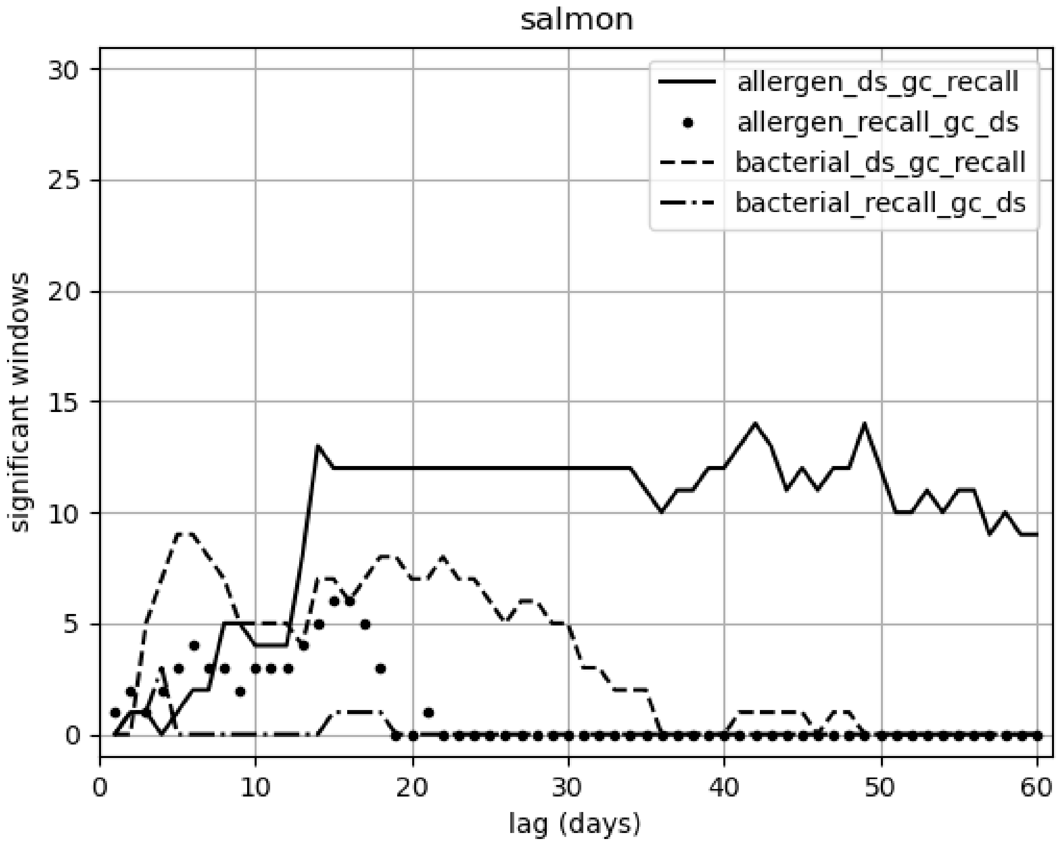
salmon
allergen_ds_gc_recall
allergen_recall_gc_ds
bacterial_ds_gc_recall
bacterial_recall_gc_ds
significant windows
lag (days)
0
5
10
15
20
25
30
0
10
20
30
40
50
60

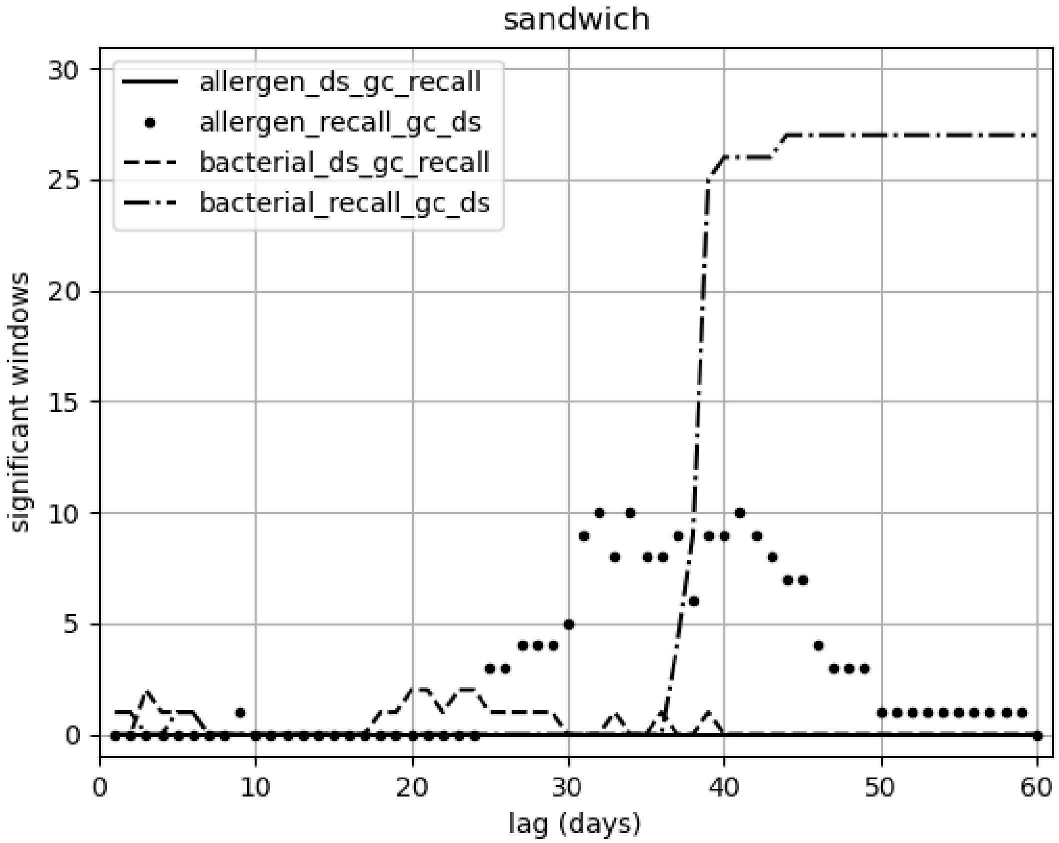
sandwich
allergen_ds_gc_recall
allergen_recall_gc_ds
bacterial_ds_gc_recall
bacterial_recall_gc_ds
significant windows
lag (days)
0
5
10
15
20
25
30
40
50
60

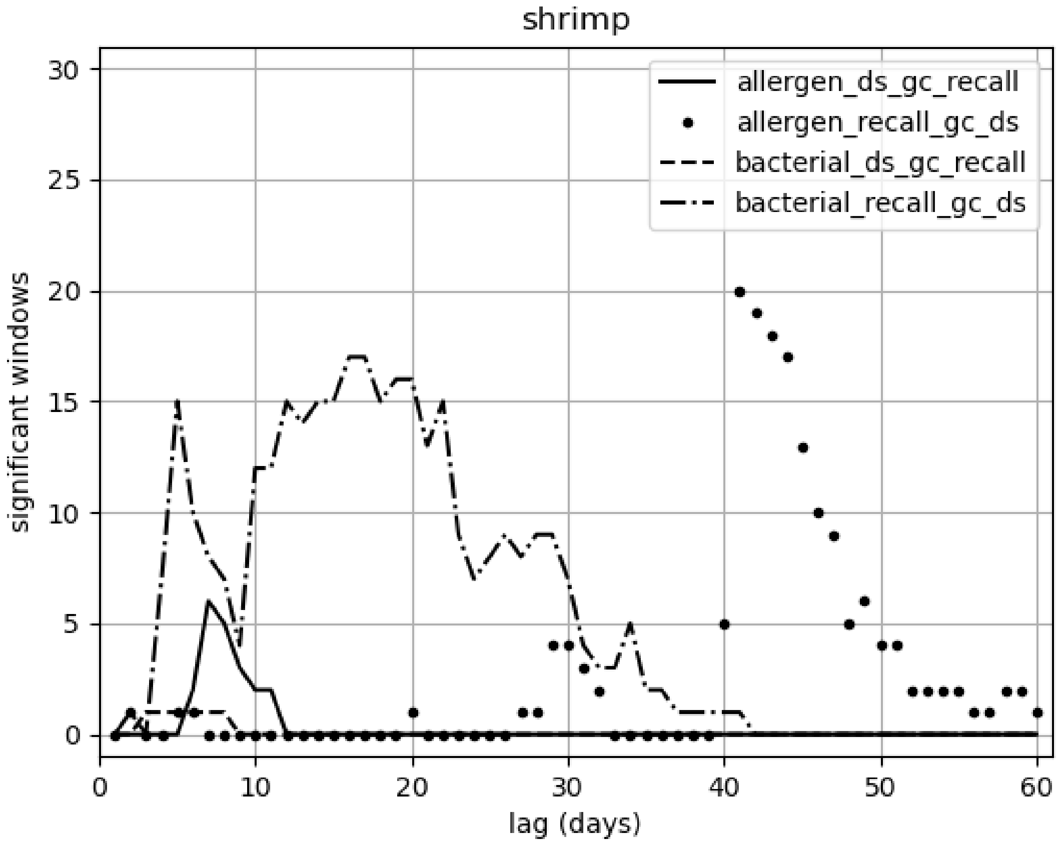
shrimp
allergen_ds_gc_recall
allergen_recall_gc_ds
bacterial_ds_gc_recall
bacterial_recall_gc_ds
significant windows
lag (days)
0
5
10
15
20
25
30
40
50
60

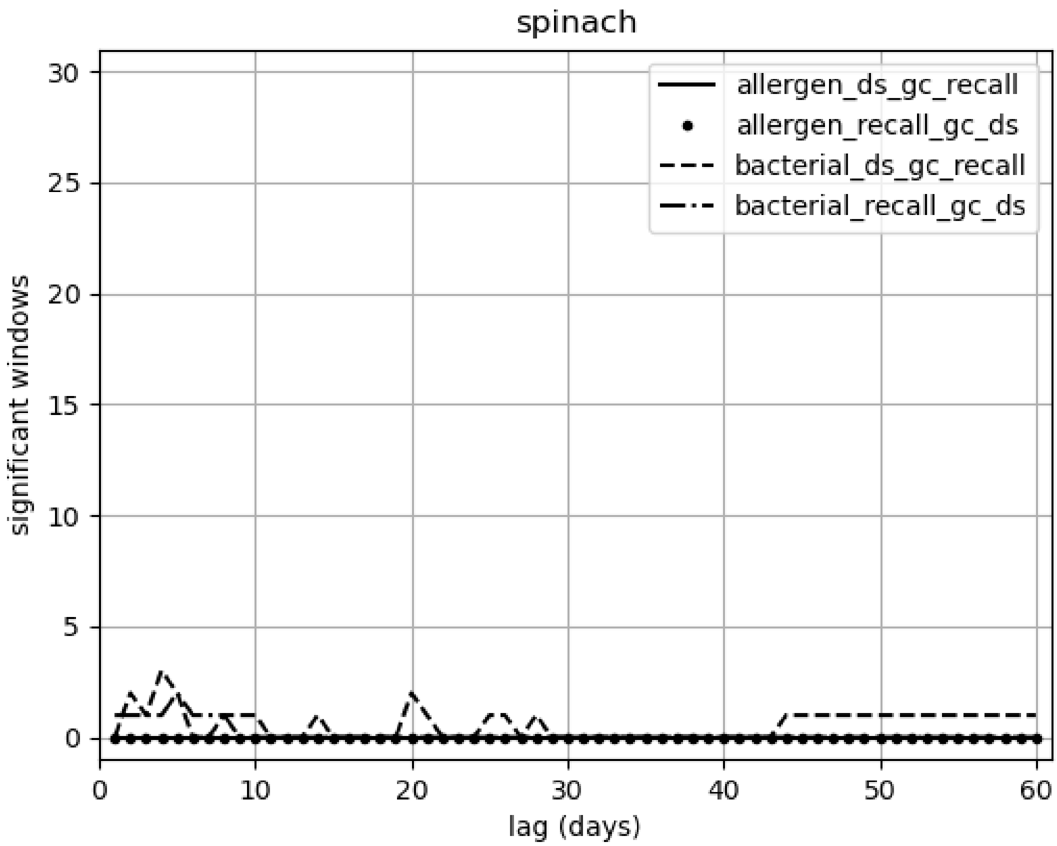

spinach
allergen_ds_gc_recall
allergen_recall_gc_ds
bacterial_ds_gc_recall
bacterial_recall_gc_ds
significant windows
lag (days)
0
5
10
15
20
25
30
40
50
60

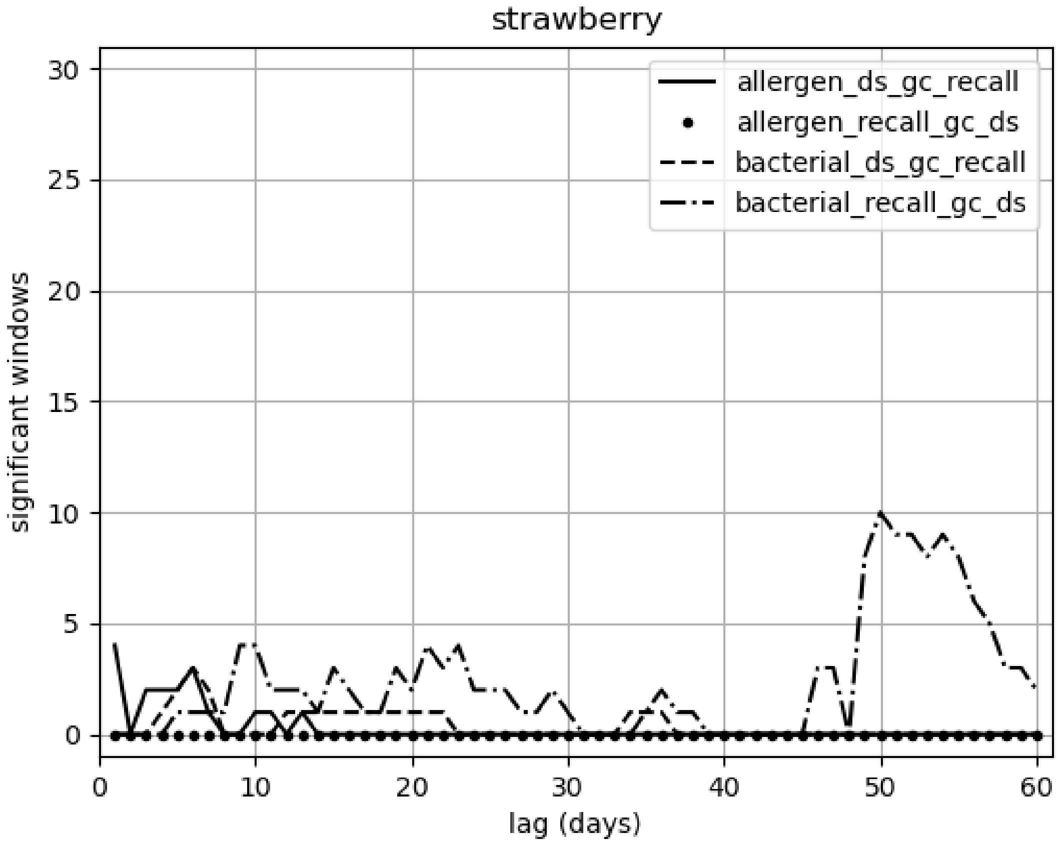

strawberry
allergen_ds_gc_recall
allergen_recall_gc_ds
bacterial_ds_gc_recall
bacterial_recall_gc_ds
significant windows
lag (days)
0
5
10
15
20
25
30
0
10
20
30
40
50
60

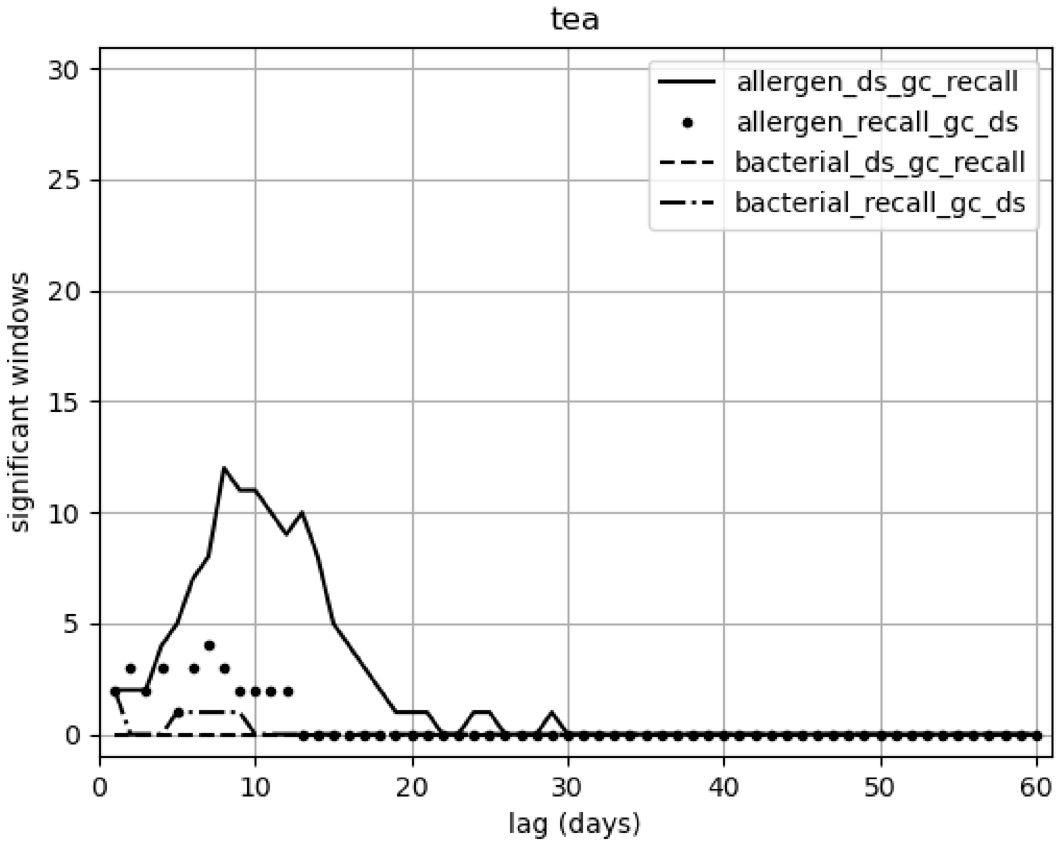

tea
allergen_ds_gc_recall
allergen_recall_gc_ds
bacterial_ds_gc_recall
bacterial_recall_gc_ds
significant windows
lag (days)
0
5
10
15
20
25
30
40
50
60

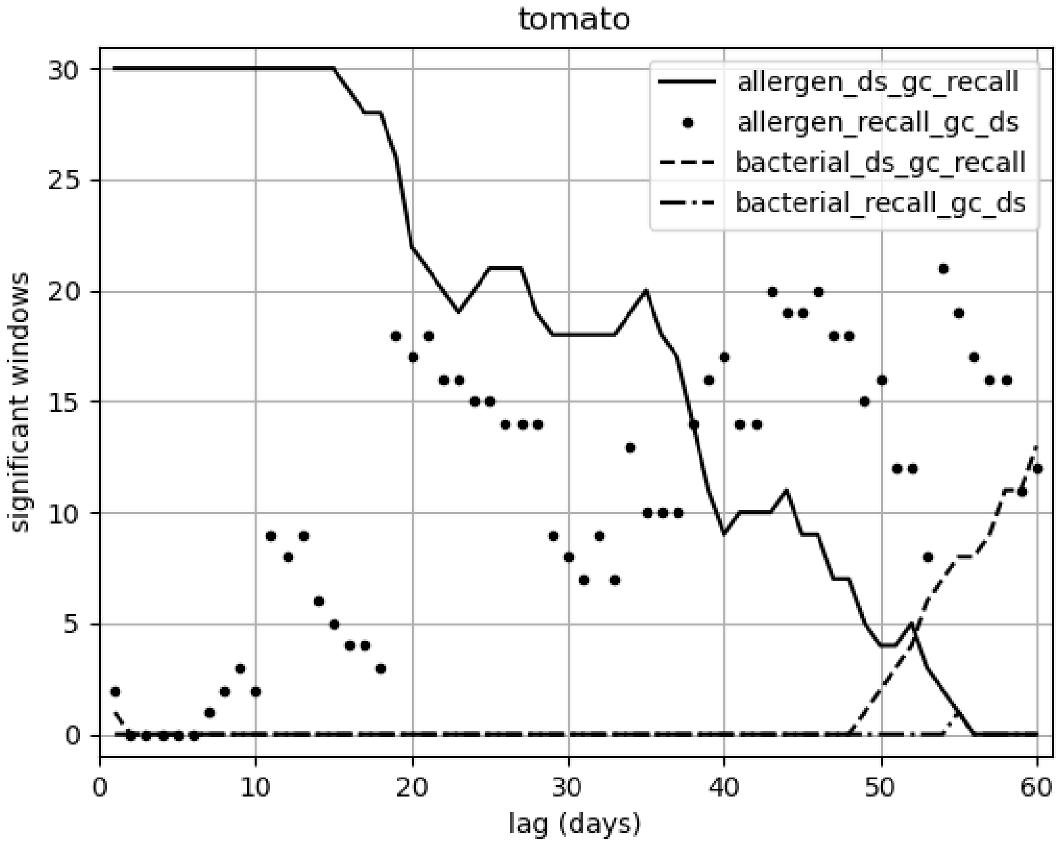
tomato
significant windows
lag (days)
allergen_ds_gc_recall
allergen_recall_gc_ds
bacterial_ds_gc_recall
bacterial_recall_gc_ds
0
10
20
30
40
50
60
0
5
10
15
20
25
30

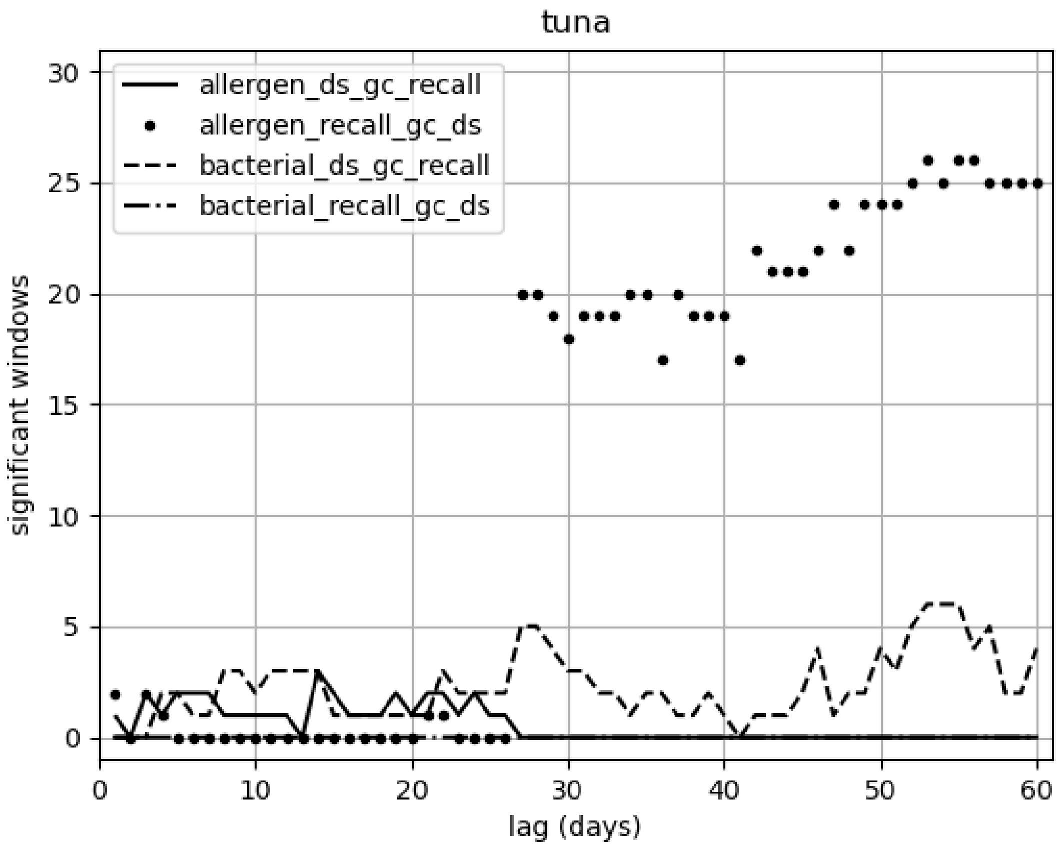
tuna
allergen_ds_gc_recall
allergen_recall_gc_ds
bacterial_ds_gc_recall
bacterial_recall_gc_ds
significant windows
lag (days)
0
5
10
15
20
25
30
0
10
20
30
40
50
60

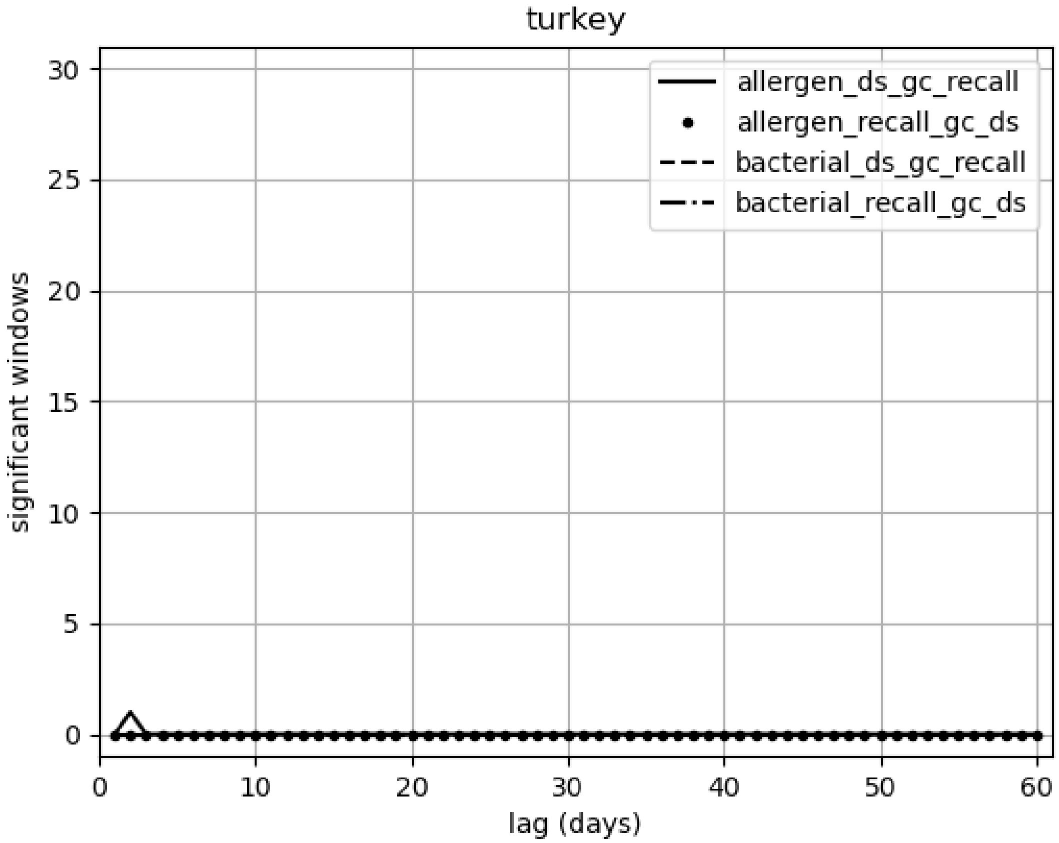

turkey
allergen_ds_gc_recall
allergen_recall_gc_ds
bacterial_ds_gc_recall
bacterial_recall_gc_ds
significant windows
lag (days)
0
5
10
15
20
25
30
0
10
20
30
40
50
60
